# Insights Into Parkinson’s Disease Genetics in African Populations: Expanded GWAS Identifies Ancestry-Specific and Cross-Population Risk Loci

**DOI:** 10.64898/2026.03.01.26347367

**Authors:** Njideka Okubadejo, Oluwadamilola O Ojo, Oladunni Abiodun, Sani Abubakar, Fatimah Abdulai, Charles Achoru, Osigwe Agabi, Uchechi Agulanna, Rufus Akinyemi, Wemimo Alaofin, Roosevelt Anyanwu, Cyril Erameh, Daniel Ezuduemoih, Abdullahi Ibrahim, Erica Ikwenu, Frank Imarhiagbe, Ismaila Ishola, Emmanuel Iwuozo, Morenikeji Komolafe, Alero Nnama, Paul Nwani, Franscisca Nwaokorie, Ernest Nwazor, Yahaya Obiabo, Nkechi Obianozie, Olanike Odeniyi, Francis Odiase, Ewere Marie Ogbimi, Adebimpe Ogunmodede, Francis Ojini, Rashidat Olanigan, Adedunni Olusanya, Chiamaka Okereke, Gerald Onwuegbuzie, Godwin Osaigbovo, Nosakhare Osemwegie, Olajumoke Oshinaike, Lukman Owolabi, Raymond Owolabi, Shyngle Oyakhire, Simon Izuchukwu Ozomma, Fadimatu Sa’Ad, Funmilola Taiwo, Kolawole Wahab, Mie Rizig, Stella Aslibekyan, Matt Kmiecik, Karl Heilbron, 23andMe Research Team, Kamalini Ghosh Galvelis, Cyrus P Zabetian, Kathryn Step, Jonathan Carr, Soraya Bardien, Pawel Lis, Lana Chahine, Naomi Louie, Alyssa O’Grady, Shivika Chandra, Marissa Dean, Elizabeth Disbrow, Deborah Hall, Vanessa Hinson, Camilla Kilbane, Scott Norris, Ashley Rawls, Ejaz Shamim, Lisa Shulman, Julia Staisch, Tao Xie, Andrew Ameri, Erin Foster, Erin Furr Stimming, Natalia Pessoa Rocha, Lietsel Jones, Lara Lange, Zih-Hua Fang, Kristin Levine, Huw Morris, Mike Nalls, Cornelis Blauwendraat, Andrew Singleton, Hampton Leonard, Mary B Makarious, Global Parkinson’s Genetics Program (GP2)

## Abstract

**Introduction:** Genome-wide association studies (GWAS) have identified over 130 risk loci for Parkinson’s disease (PD), yet the majority derive from studies performed in European ancestry populations. African (AFR) and African admixed (AAC) ancestry individuals remain underrepresented in PD genetics research, limiting our understanding of ancestry-specific genetic architecture and the generalizability of known risk factors.

**Methods:** We conducted GWAS in AFR and AAC populations by integrating individual-level genotype data from the Global Parkinson’s Genetics Program (GP2) with summary statistics from 23andMe Research Institute and the Million Veterans Program. The combined dataset included 3,975 cases and 319,883 controls, representing a 64% increase in total sample size compared with prior analyses. We performed separate GWAS for AFR and AAC cohorts as well as a combined AFR/AAC meta-analysis.

**Results:** The intronic *GBA1* variant rs3115534 was the most significant association across all analyses, reaching genome-wide significance in AAC individuals for the first time. In the AFR-only analysis, five loci achieved genome-wide significance: *GBA1* (rs3115534), the *SNCA* signal previously reported in European ancestry GWAS (rs356182), a new protein-coding association at *LRRK2* (rs72546327, p.T1410M), a non-coding *RPL10P13* variant (rs12302417), and a novel signal on chromosome 16 (rs113244182). The combined AFR/AAC meta-analysis identified four genome-wide significant associations at *GBA1* (rs3115534), *SNCA* (rs356182), *SCARB2* (rs11547135), and *LRRK2* (rs139283662, which is in LD with p.T1410M).

**Conclusions:** This study reports the largest GWAS of PD in AFR and AAC populations to date. Our findings confirm trans-ancestry risk loci (*GBA1* and *SCARB2*) and identify an ancestry-enriched coding variant at *LRRK2*. This convergence of evidence around genes involved in glucocerebrosidase (GCase) trafficking and alpha-synuclein clearance supports current therapeutic strategies targeting this pathway and provides critical targets for developing precision medicine in African ancestry populations. Importantly, the identification of a novel association between a *LRRK2* coding variant with disease in the AFR and AAC populations opens up a traditionally underrepresented population for ongoing *LRRK2* targeted trials. Furthermore, the identification of novel ancestry-specific loci, including those that are directly relevant to current therapeutic deployment, underlines the importance of understanding the basis of disease in all populations.

## INTRODUCTION

Parkinson’s disease (PD) presents a rapidly growing global health challenge, with prevalence expected to nearly triple from 6.1 million in 2016 to over 17.5 million by 2040^1,2^. Genome-wide association studies (GWAS) have successfully identified over 130 risk loci in populations of European ancestry^3^, while studies in East Asian ancestry populations have identified two population-specific signals^4^, and multi-ancestry analyses have identified additional novel loci^5,6,7,8^. However, the extent to which European-derived findings can be applied to other ancestries remains unknown^9,10^.

This knowledge gap is particularly pronounced for African and African admixed populations, where epidemiological patterns have been reported to differ from European ancestry populations. Studies in sub-Saharan Africa report lower prevalence rates (30 to 60 per 100,000) compared to North Africa and Europe, and argue for distinct genetic profiles from those seen in Northern European ancestry groups ^11–14,15^. Additionally, clinical presentations in the region are often complicated by delayed diagnosis and phenotypic variation^16,17,18–20^. Consequently, a majority of genomic diversity in PD remains under-investigated, obscuring ancestry-specific mechanisms that could drive the development of more universal therapeutics.

Understanding the genetic basis of disease across diverse global groups is important^21–27^. As medicine moves toward the implementation of precision therapeutic approaches and genetically-targeted therapeutics, understanding the genetic architecture across populations becomes an imperative. It is important to perform genetic research in ancestrally diverse groups, not only to support health equity, but also to harness the scientific power of comparative genetics across ancestral groups. African genomes contain the highest levels of genetic variability and the shortest linkage disequilibrium (LD) blocks of any human population. These genomic features, particularly in comparison with other ancestral groups, facilitate high-resolution fine-mapping, allowing for the differentiation of causal variants from linked markers^28,29^.

In our previous study^30^, analysis of African-ancestry-specific variation identified a novel common risk factor at the *GBA1* locus. Unlike European *GBA1* associations, which are frequently driven by coding variants, this signal is attributed to an intronic variant (rs3115534-G) located within a key intronic branchpoint sequence. Using full-length RNA transcript sequencing and CRISPR editing, we determined that this sequence alteration disrupts splicing, leading to partial intron 8 retention. This retention prevents the translation of functional protein, resulting in a dose-dependent reduction of glucocerebrosidase activity (GCase) in risk variant carriers^31^. The variant has also been associated with increased REM sleep behavior disorder symptoms in Nigerian individuals with PD^32^. These data demonstrate an RNA-based mechanism for lysosomal dysfunction distinct from previously described coding mutations, highlighting a therapeutic target identifiable only through the study of diverse genetic architectures. Importantly, this work shows that this variant is not only a risk factor for disease, but that it also leads to an earlier disease onset of approximately 3 years per risk allele^30^.

Building on the foundational work of the Global Parkinson’s Genetics Program (GP2) and the study by Rizig et al.^30^, this study presents an expanded genome-wide assessment incorporating data from three datasets: GP2, 23andMe Research Institute, and the Million Veteran Program (MVP). The current analysis includes 3,975 cases and 319,883 controls, representing a 64% increase in sample size. Of these, 2,976 cases (74.87%) were genotyped using the NeuroBooster array (NBA) and processed through the standardized GP2 pipeline. This expanded cohort provides increased statistical power to characterize genetic risk factors in African and African admixed ancestries.

## METHODS

### Study Design

An overview of the study design is presented in **Figure 1**. This analysis integrated data from three primary sources: individual-level genotypes generated through GP2, as well as aggregated genotype GWAS summary statistics from 23andMe Research Institute, and the Million Veteran Program (MVP). Case-control counts per data source are outlined in **Table 1**. The majority of PD cases contributed through African recruitment efforts originate from West Africa, particularly Nigeria, and therefore do not represent the genetic diversity across the African continent.

**Figure 1.**
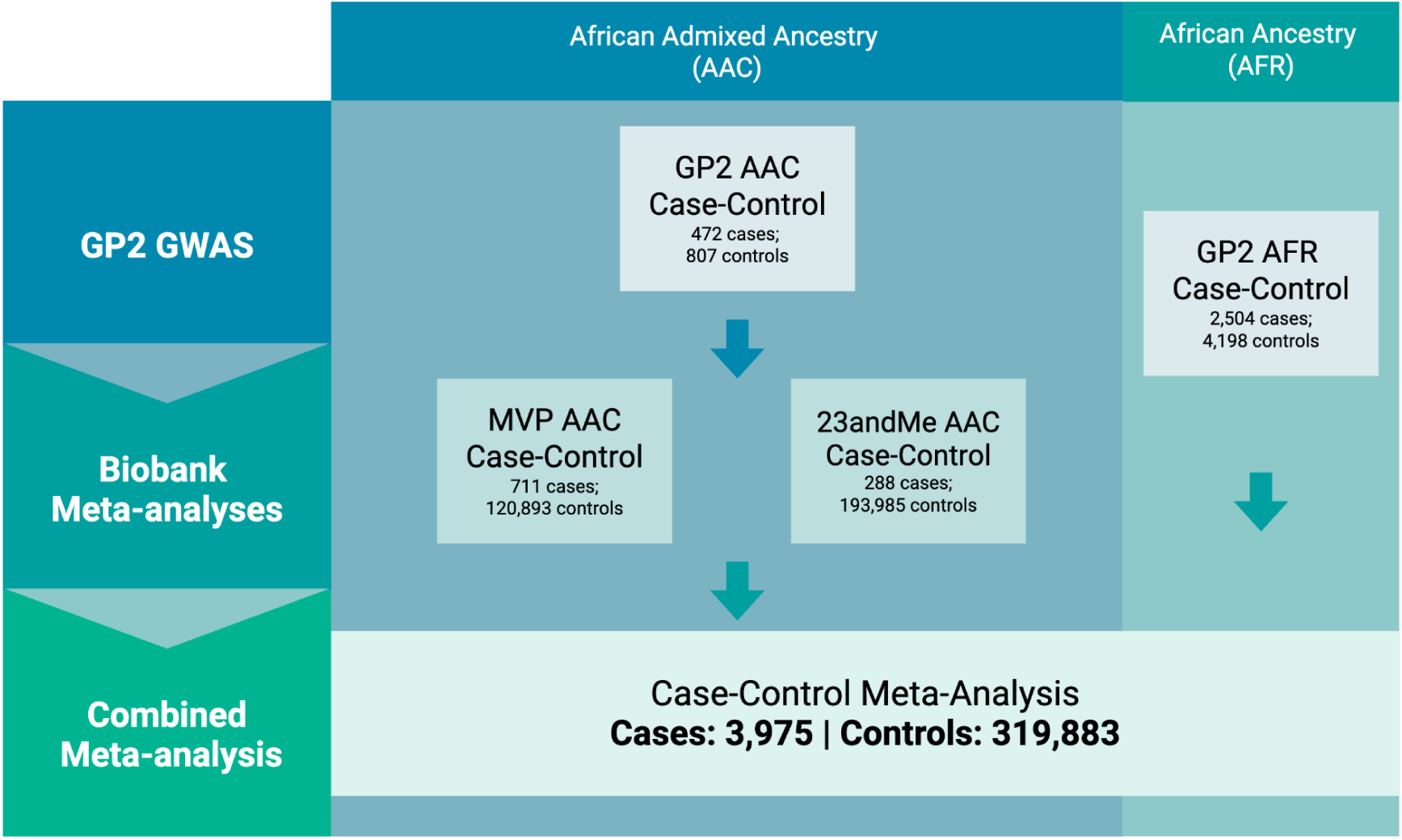
Analysis Workflow Diagram. | GP2: Global Parkinson’s Genetics Program; MVP: Million Veteran Program; AAC: African admixed ancestry; AFR: African ancestry

**Table 1.** Cohort characteristics and case-control composition across datasets. | Aggregated genotype summary statistics were provided by 23andMe and MVP GP2: Global Parkinson’s Genetics Program; MVP: Million Veteran Program; AAC: African admixed ancestry; AFR: African ancestry; PC: Principle component

| Cohort | Ancestry | Cases | Controls | % Female cases | % Female controls | Array type | Imputation Summary | Covariates used for Cohort-level GWAS |
| --- | --- | --- | --- | --- | --- | --- | --- | --- |
| GP2 AAC R11 | African admixed (AAC) | 472 | 807 | 39.407 | 63.817 | NeuroBooster Array | TOPMed R3, Freeze 10; Michigan server for rs3115534 | Sex, PC1-20 |
| GP2 AFR R11 | African (AFR) | 2,504 | 4,198 | 30.351 | 48.428 | NeuroBooster Array | TOPMed R3, Freeze 10; Michigan server for rs3115534 | Sex, PC1-20 |
| 23andMe | African admixed (AAC) | 288 | 193,985 | NA | NA | Omni Express & GSA & 550k | Human Reference Consortium (HRC) panel and custom 23andMe reference panel | Summary statistics |
| MVP | African admixed (AAC) | 711 | 120,893 | NA | NA | Custom Affymetrix Axiom array (MVP 1.0 Genotyping Array) | Hybrid panel: African Genome Resources and 1000 Genomes Phase 3 | Summary statistics |

GP2 is an international initiative that combines ancestry-diverse genetic and clinical data from PD and related dementia cohorts worldwide to accelerate discovery and enable equitable research with the goal of advancing precision medicine^23^. For all GP2 cohorts, PD diagnoses were made according to the United Kingdom Parkinson’s Disease Society Brain Bank criteria, with the exception of the criterion limiting the number of affected relatives^33^. Ethical approval for participation in genetic research was obtained from the relevant institutional committees, and all individuals provided written informed consent.

Neurologists or qualified study staff performed standardized examinations on all PD participants to document motor and non-motor symptoms. Control participants were evaluated for general neurological health, and individuals exhibiting signs of neurodegenerative disease were excluded from the control group.

23andMe is non-profit medical research organization that uses consented participant genotype and phenotype data to perform population-scale genetic research across a wide range of traits and diseases. For the 23andMe dataset, PD status was self-reported, with summary statistics provided under a research collaboration with 23andMe, Inc. All participants provided informed consent and volunteered to participate in the research online, under a protocol approved by the external AAHRPP-accredited Salus IRB (https://www.versiticlinicaltrials.org/salusirb). Additional details on recruitment and phenotype definitions are provided elsewhere^30^.

The MVP is a national cohort launched in 2011 to study the contributions of genetics, lifestyle, and military exposures to health and disease among US Veterans. For the MVP dataset, PD summary statistics from the Million Veteran’s Program (MVP) were downloaded from dbGAP (accession number: phs002453.v1.p1; analysis accession: pha010400.1). Details on recruitment and phenotype definitions can be found in the dbGaP documents, as well as elsewhere^34^. The African American PD analysis included 121,604 Veterans (711 cases, 120,893 controls) defined by PheCode 332 (Parkinson’s disease). Cases were defined as Veterans with 2 or more instances of PheCode 332-mapped ICD-9 or ICD-10 codes extracted from the VA electronic health record through September 2019. Controls had no recorded instances of PheCode 332-mapped codes and no ICD-10 codes for G20, G21, G22, G23, G24, or G25.

### Cohort Quality Control and Ancestry

### Global Parkinson’s Genetics Program Genotype Data

All GP2 samples (release 11; DOI 10.5281/zenodo.17753486) were genotyped on the Illumina NeuroBooster Array (version 1.0; Illumina, San Diego, CA, USA), which includes 1,914,935 backbone variants along with ancestry-informative markers, IBD markers, X-chromosome SNPs for sex inference, and 96,517 custom variants^35^. Raw array data were processed using the GenoTools pipeline (available at https://github.com/GP2code/GenoTools; ^36^, which incorporates a machine-learning framework for ancestry prediction and relatedness pruning.

Ancestry estimation was performed using a unified reference panel derived from three large resources: the 1000 Genomes Project^37^, the Human Genome Diversity Project^38^, and a dataset focused on the Ashkenazi Jewish population^39^. The combined reference set included 4,508 individuals representing African (n=703), African admixed (n=190), European (n=534), Finnish (n=99), Latin American (n=490), East Asian (n=585), South Asian (n=601), Central Asian (n=183), Middle Eastern (n=152), and Ashkenazi Jewish (n=471) populations. Reference variants were filtered to remove palindromic SNPs, MAF <5%, call rate <99%, and Hardy-Weinberg equilibrium (HWE) P<1E-4. GP2 genotypes were harmonized to this panel, with missing data imputed using mean allele dosages. Principal components (PCs) were derived from overlapping SNPs and transformed with UMAP to improve population predictions. A linear support vector machine trained using an 80:20 train:test split achieved >0.95 balanced accuracy in 5-fold cross-validation, and was used to assign ancestry labels to GP2 samples.

Sample-level QC excluded individuals with call rate <95%, discordance between genetic and reported sex, excess heterozygosity (|F|>0.25), or relatedness exceeding 0.088 kinship (IBD = 17.6%; approximately 2nd degree relatives). SNP-level QC removed variants with HWE P<1E-4 in controls, or with differential missingness by case-control status or haplotype at P<=1E-4.

Phased datasets were submitted to the TOPMed Imputation Server^40^ using the TOPMed r3 reference panel, which includes 133,597 reference samples and 445 million variants across autosomes and the X chromosome^41,42^. Post-imputation QC retained variants with imputation quality R^2^ >=0.3 for all downstream genome-wide analyses. Although the TOPMed r3 reference panel provides dense genomic coverage, it does not impute several ancestry-enriched risk variants that were reliably imputed in the TOPMed r2 reference panel, including rs3115534 located within the *GBA1* locus. To recover genotypes for this variant, we performed targeted imputation. Specifically, imputation of chromosome 1 was conducted in the GP2 AAC and AFR datasets using the high-coverage 30x 1000 Genomes Phase 3 reference panel provided by the Michigan Imputation Server^41^. Following imputation, genotypes corresponding to rs3115534 were merged back into the primary TOPMed-imputed genotype files to restore variant-level completeness. This allowed the inclusion of the variant in downstream analyses without compromising the broader imputation framework. R2 values per ancestry for all variants nominated in this study can be found in **Supplementary Table 6**.

### 23andMe Summary Statistics

Participants were genotyped on one of five Illumina-based platforms (v1-v5), ranging from ∼560,000 to ∼950,000 variants, with samples failing to achieve 98.5% call rate re-genotyped. Ancestry assignment utilized a proprietary ancestry composition algorithm that analyzed local ancestry by partitioning phased genotype data into windows of approximately 300 SNPs. A support vector machine classified individual haplotypes into one of 45 reference populations derived from the African Genetics Project, Human Genome Diversity Project, HapMap, and 1000 Genomes Project, among others. Classifications were refined using a hidden Markov model and calibrated with simulated admixed individuals. Reference populations were grouped into six higher-level ancestries: African American, East Asian, European, Latin American, South Asian, and Native American. To distinguish between African American and Latin American individuals, who share overlapping continental ancestry proportions, a logistic classifier was trained using length distributions of ancestry-specific genomic segments, which differ due to distinct admixture histories.

Phasing was performed using SHAPEIT4 with platform-specific reference panels built from 200,000 high-quality African American participants. Imputation utilized a hybrid approach combining the Human Reference Consortium panel (27,165 samples, 39 million variants) and a 23andMe-specific panel (12,217 samples from internal and external whole-genome sequencing datasets), implemented in Beagle 5. Relatedness filtering excluded individuals sharing >700 cM identity-by-descent, approximately equivalent to first-cousin relationships, with cases preferentially retained over controls. Additional details on quality control, imputation, and ancestry prediction are provided elsewhere^30^.

### MVP Summary Statistics

MVP participants were genotyped using a custom Affymetrix Axiom biobank array (MVP 1.0 Genotyping Array). Sample-level quality control excluded duplicate samples, samples with observed heterozygosity exceeding expected levels, missing genotype call rate >2.5%, or sex discordance between genetic and phenotypic data. Variant-level QC removed probes with missingness >20%, monomorphic variants, and those with Hardy-Weinberg equilibrium P<1E-6 in either the overall cohort or within major ancestry groups.

Ancestry assignment utilized genetically inferred ancestry (GIA), as elaborated on in the MVP gwPheWAS publication^43^. Population-specific principal components were computed using EIGENSOFT v.6. Genetic imputation was performed using a hybrid imputation panel comprising the African Genome Resources panel and 1000 Genomes Phase 3 reference, implemented with SHAPEIT4 (v4.1.3) and Minimac4.

Additional details on quality control, imputation, and ancestry prediction are provided elsewhere^34^.

### GWAS, Meta-analyses, and follow-up analyses

For the 23andMe and MVP datasets, only minimal quality-control filters were applied, primarily to address outlier frequency values and variants reporting an effect estimate of zero. As individual-level data were not available, ancestry for both datasets was inferred from study-specific documents as well as country-of-origin information and classified as African admixed (AAC).

For the GP2 datasets, GWAS were conducted using Plink2^44^, correcting for PCs and genetically-determined sex. Related individuals (2nd degree or closer) were removed prior to analysis.

For all datasets, all aggregated genotype summary statistics were filtered for minor allele frequency (MAF) > 1% specific to each study, except for the *GBA1*, *SNCA*, and *LRRK2* regions (gene ±250kb) in order to capture rare variants of potential interest. Fixed-effect and random-effects meta-analyses were carried out per ancestry and per case ascertainment criteria (clinical case-control or biobank) as described, and then finally for the full joint analysis using Plink1.9^45^. Significant independent loci were determined by the genome-wide significance threshold of P<5E-8 and by using a linkage disequilibrium (LD) clumping threshold of r2 > 0.6 within 500-kb windows, as per gwaslab default thresholds^46^.

For GWAS loci follow-up, we leveraged whole blood expression quantitative trait locus (eQTL) summary statistics from Kachuri and colleagues, which were derived from African American, Puerto Rican, and Mexican individuals (Kachuri et al. 2023). Specifically, we used the AFRHp5 subset, comprising individuals with >50% African ancestry, to ensure appropriate ancestral matching with our African GWAS cohort. Additionally, we used LocusCompare to assess correlations between GWAS and between GWAS and eQTL studies.

## RESULTS

We conducted the largest PD GWAS to date in AFR and AAC populations, using individual-level genotype data from the Global Parkinson’s Genetics Program (GP2) and meta-analyzing these with aggregated genotype summary statistics from 23andMe and the Million Veteran Program. The combined dataset included 3,975 cases and 319,883 controls. A summary of all GWAS performed is shown in **Figure 1** and all genomic inflation factors (lambdas) for all studies and meta-analyses are in **Supplementary Table 1**.

### African Admixed ancestry Parkinson’s disease Meta-Analysis

We first evaluated PD risk in AAC individuals by performing a meta-analysis of GWAS summary statistics generated from GP2, 23andMe, and MVP (**Figure 2**). Earlier work using smaller AAC datasets had not identified any genome-wide significant associations^30^. With the expanded sample size in the present study, we now observed a clear genome-wide significant signal at the *GBA1* locus. The lead variant, rs3115534 (chr1:155235878:G:T), reached genome-wide significance in the AAC meta-analysis (beta = -0.33, SE = 5.47E-02; p = 1.36E-9), marking the first time this locus has surpassed the threshold in African admixed individuals. This effect size is similar to the previously reported -0.458 (SE = 0.059) by Rizig and colleagues in African populations. No additional loci reached genome-wide significance in the AAC-only analysis.

**Figure 2.**
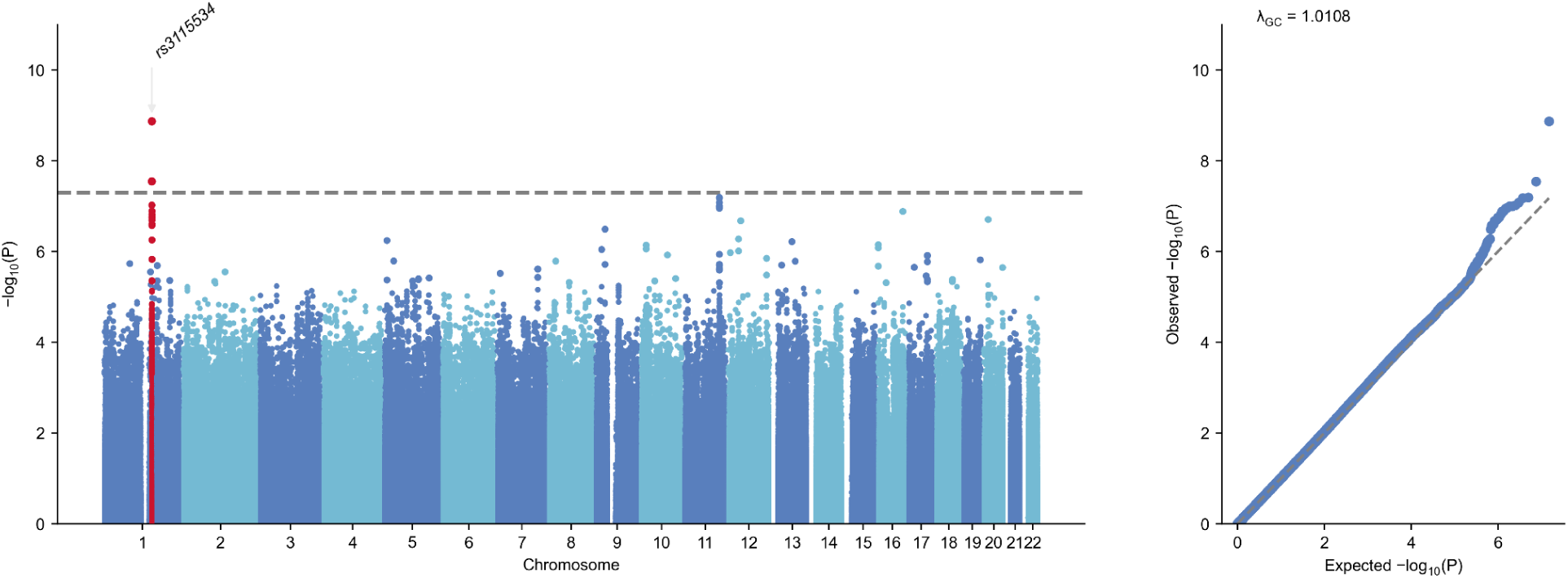
AAC GWAS Meta-analysis Manhattan and QQ Plot. Includes GP2 AAC, 23andMe, MVP

### African Ancestry Parkinson’s disease GWAS

Next, we next conducted a GWAS in individuals of AFR ancestry using GP2 individual-level genotype data (**Figure 3**). With a considerably larger sample size than in prior analyses, five loci surpassed genome-wide significance. As expected from previous findings, the strongest association was detected at *GBA1*, with the functional branch site variant, rs3115534, showing a highly significant association with PD risk (beta = -0.55, SE = 0.042; p = 1.2E-37). Four additional independent loci were identified at genome-wide significance. The intronic 3’ *SNCA* variant rs356182, which is an established risk variant for PD in Northern European populations and other ancestries, showed a strong association with PD in the AFR analysis (beta = -0.28; SE = 0.0039; p = 4.33E-13). A *LRRK2* missense variant, p.T1410M (rs72546327) was associated with risk for disease (beta = 0.55; SE = 0.0097; p = 1.72E-8). A non-coding variant, rs12302417, within the *RPL10P13* pseudogene also showed association with disease (beta = -0.32; SE =0.06; p =4.49E-8). Lastly, the intergenic variant rs113244182 on chromosome 16 was associated with PD risk (beta = -0.80; SE = 0.14; p = 1.67E-8). See **Supplementary Figures 3-9** for Forest plots and **Supplementary Figures 10-27** for LocusZoom plots. These findings reflect the greater power of the expanded African dataset and highlight multiple loci contributing to PD susceptibility in individuals of African ancestry.

**Figure 3.**
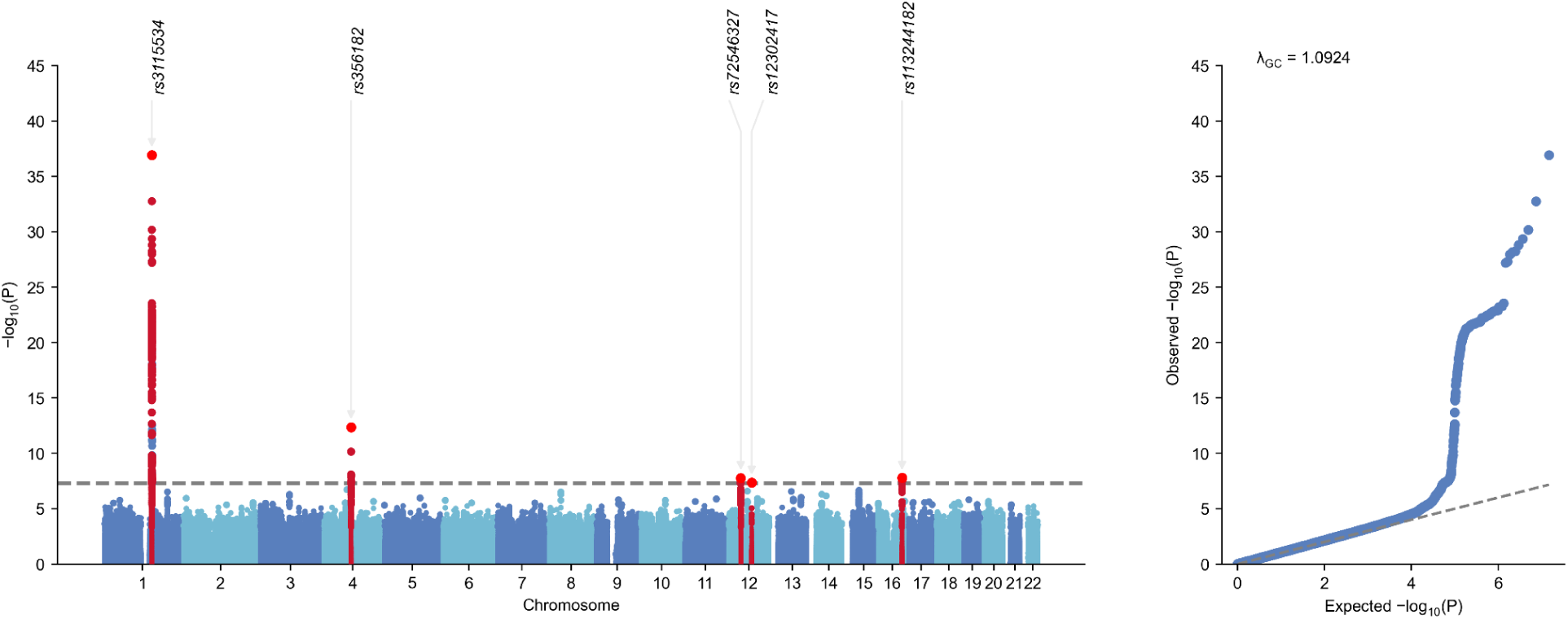
AFR GWAS Manhattan and QQ Plot. Includes GP2 AFR

### Combined African Admixed and African Meta-Analysis

To maximize power and assess shared signals across AFR and AAC groups, we performed a combined meta-analysis incorporating GP2 AFR, GP2 AAC, 23andMe, and MVP summary statistics (**Figure 4**). Four loci achieved genome-wide significance in this analysis. The most significant association remained rs3115534 at *GBA1* (beta = -0.47, SE = 0.0337; p = 1.53E-43), reinforcing *GBA1* as the leading common PD risk signal in African ancestry populations. Three additional loci were identified at genome-wide significance: rs11547135 at *SCARB2* (beta = 0.18, SE = 0.0330; p = 3.51E-8), rs356182 at *SNCA* (beta = -0.24, SE =0.0295; p = 8.38E-16), and rs139283662 in the *LRRK2* region (beta = 0.63, SE = 0.1098; p = 1.25E-8), see **Supplementary Figures 3-9** for Forest plots and **Supplementary Figures 10-27** for LocusZoom plots. Notably, rs139283662 represents the same underlying genetic signal at *LRRK2* as rs72546327, the lead variant identified in the African ancestry analysis above. Examination of linkage disequilibrium shows that rs72546327 lies approximately 0.28 Mb from the variant rs139283662 within the intron of *SLC2A13* (beta = 0.63; p = 1.25E-8) detected in the AFR/AAC meta-analysis, and the two variants are in LD in African populations (D′ = 1.0; R² = 0.41; https://ldlink.nih.gov/?tab=ldpair), thus it is likely that the rs139283662 signal is tagging the rarer *LRRK2* coding variant rs72546327 (p.T1410M). Together, the results of the combined meta-analysis underscore both shared and ancestry-specific contributors to PD risk across African and African admixed populations.

**Figure 4.**
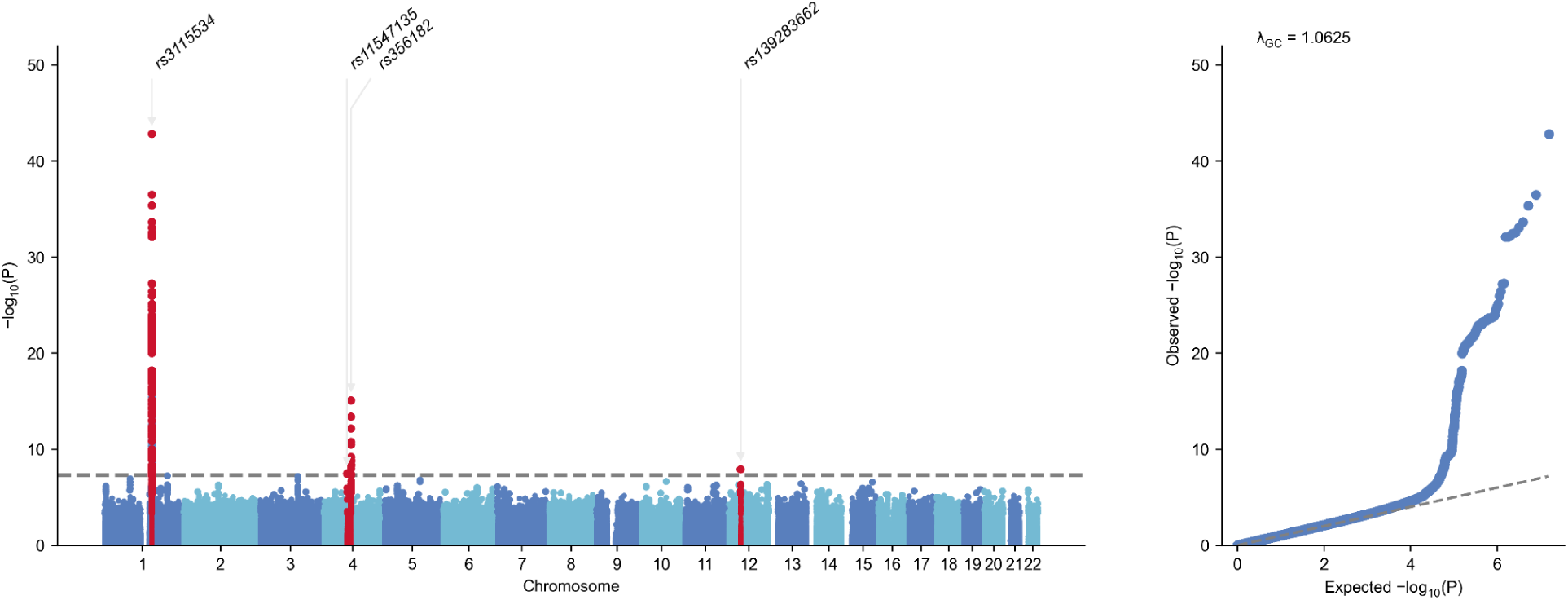
AFR/AAC GWAS Meta-analysis Manhattan and QQ Plot. Includes GP2 AFR, GP2 AAC, MVP, 23andMe

### Investigation of African Admixed and African GWAS hits

Next, we investigated the identified AFR and AAC GWAS hits to determine whether the signals that are physically close (for example, near *LRRK2*) represent independent disease associations, or simply a single signal tagged by different variants. In total, seven unique GWAS loci were identified across the three analyses. As previously mentioned, out of these seven GWAS loci, five have already been identified in previous PD GWAS of other ancestries (**Table 2**). For the already identified GWAS loci (*GBA1*, *SNCA*, *SCARB2/FAM47E*, *LRRK2*, *SLC2A13*), we aimed to assess whether the identified AFR and AAC GWAS hits represent the same signal as previous GWAS or distinct signals. Given the prior identification of the *GBA1* locus (rs3115534) in AFR and the functional follow-up, we consider this locus resolved.

**Table 2.** Summary of genome-wide loci nominated in study. SNP IDs provided in hg38 AAC: African admixed ancestry; AFR: African ancestry; SE: Standard Error

| Analysis | SNP ID | Effect Allele | rsID | BETA | SE | P | Close to Known Locus? |
| --- | --- | --- | --- | --- | --- | --- | --- |
| AAC Meta-analysis | chr1:155235878:G:T | T | rs3115534 | -0.3317 | 0.0547 | 1.36E-09 | Yes; *GBA1* |
| AFR | chr1:155235878:G:T | T | rs3115534 | -0.54839 | 0.04276 | 1.20E-37 | Yes; *GBA1* |
|  | chr4:89704960:G:A | A | rs356182 | -0.28492 | 0.03933 | 4.33E-13 | Yes; *SNCA* |
|  | chr12:40309145:C:T | T | rs72546327 | 0.54691 | 0.097 | 1.72E-08 | Yes; *LRRK2* |
|  | chr12:75689052:G:A | A | rs12302417 | -0.32457 | 0.05933 | 4.49E-08 | No; novel (*RPL10P13*) |
|  | chr16:76969646:G:C | C | rs113244182 | -0.79848 | 0.14149 | 1.67E-08 | No; novel |
| Combined AFR/AAC Meta-analysis | chr1:155235878:G:T | T | rs3115534 | -0.4662 | 0.0337 | 1.53E-43 | Yes; *GBA1* |
|  | chr4:76213633:C:T | T | rs11547135 | 0.1822 | 0.033 | 3.51E-08 | Yes; *SCARB2/FAM4E* |
|  | chr4:89704960:G:A | A | rs356182 | -0.2373 | 0.0295 | 8.38E-16 | Yes; *SNCA* |
|  | chr12:40027276:T:C | T | rs139283662 | 0.6251 | 0.1098 | 1.25E-08 | Yes; *LRRK2* |

*SNCA* has been associated on many levels with PD, Lewy body pathology, and genetically with rare causal variants and common variants through GWAS since 2009. Given that the index variant (rs356182) is identical to the European GWAS main signal^3^ and Latino GWAS^7,8^, and the overlapping LocusZoom figures (**Supplementary Figure 2**) we consider that the functional variant underlying this signal is identical across ancestries. The consistent identification of this variant as the lead signal at this locus across multiple studies and diverse ancestry groups strongly suggests that this variant is the causal variant driving disease risk at this locus.

The *SCARB2* signal was first identified as a PD risk locus in European ancestry in 2014^47^ and in the most recent European GWAS two independent signals were identified in this locus (rs13117238 and rs28628748)^3^. Of interest, *SCARB2* encodes LIMP-2, the scavenger receptor responsible for trafficking GCase (encoded by *GBA1*) from the endoplasmic reticulum to the lysosome^48^. We compared the GWAS signals across Europeans and Africans, and identified that the signal identified in the combined meta-analysis is identical to the European *SCARB2* signal (**Supplementary Figure 2**).

Finally, we also observed a significant association at *LRRK2* driven by the missense p.T1410M variant rs72546327 (beta = 0.55, SE = 0.097; p = 1.72E-8), a signal that remained nominally associated in the full combined meta-analysis (beta = 0.40, SE = 0.080; p = 6.51E-07). The case-control frequencies in GP2 data are 0.047 in AFR cases, 0.027 in AFR controls and 0.015 in AAC cases, 0.022 in AAC controls (**Supplementary Table 2**). *LRRK2* is a gene with well-known ancestral heterogeneity, with population-enriched pathogenic and susceptibility variants. For example, the globally distributed pathogenic mutation p.G2019S resides on a common haplotype of likely Middle Eastern origin, whereas the risk alleles p.G2385R and p.R1628P occur almost exclusively in Asian populations^26,49^.

Mechanistically, disease linked *LRRK2* mutations have been linked to perturbation of Lrrk2 kinase and GTPase activities, which are influenced by interactions with cellular membranes, microtubule filaments, and 14-3-3 proteins, as well as by upstream effectors such as Rab29 and *VPS35*^50^. No eQTL signals were identified to correlate with this GWAS signal (**Supplementary Figure 1**), suggesting the association may operate through mechanisms beyond gene expression regulation in blood.

Interestingly, Thr1410 lies at the C-terminal end of the Roc Switch II helix (1396–1409), positioned within a compact but structurally critical hydrophilic interface that links COR-A and COR-B. The Thr1410 side-chain hydroxyl forms a hydrogen bond with Lys1336, while its backbone interacts with Tyr1415 and Arg1441 (**Figure 5**). Arg1441 extends this network by hydrogen-bonding to the backbone of Trp1791 in COR-B, thereby physically coupling the Roc Switch II region to the COR-B lobe. Lys1336 further stabilises the interface through electrostatic interactions with Trp1393 and Glu1578 of COR-A. Arg1412 also reinforces packing in this region: its guanidinium group hydrogen-bonds to the backbone of Arg1334, and its aliphatic chain packs against Val1518 of COR-A. Taken together, these interactions position Thr1410 as a potentially important structural node within the Roc–COR interface, helping maintain communication between the Roc domain and adjacent COR subdomains. The substitution of methionine likely destabilizes this interface by introducing a bulkier, hydrophobic residue that cannot support the native hydrogen-bond network. Such disruption may propagate through the Switch II motif, potentially shifting its conformation in a manner reminiscent of GDP/GTP-dependent structural transitions.

**Figure 5.**
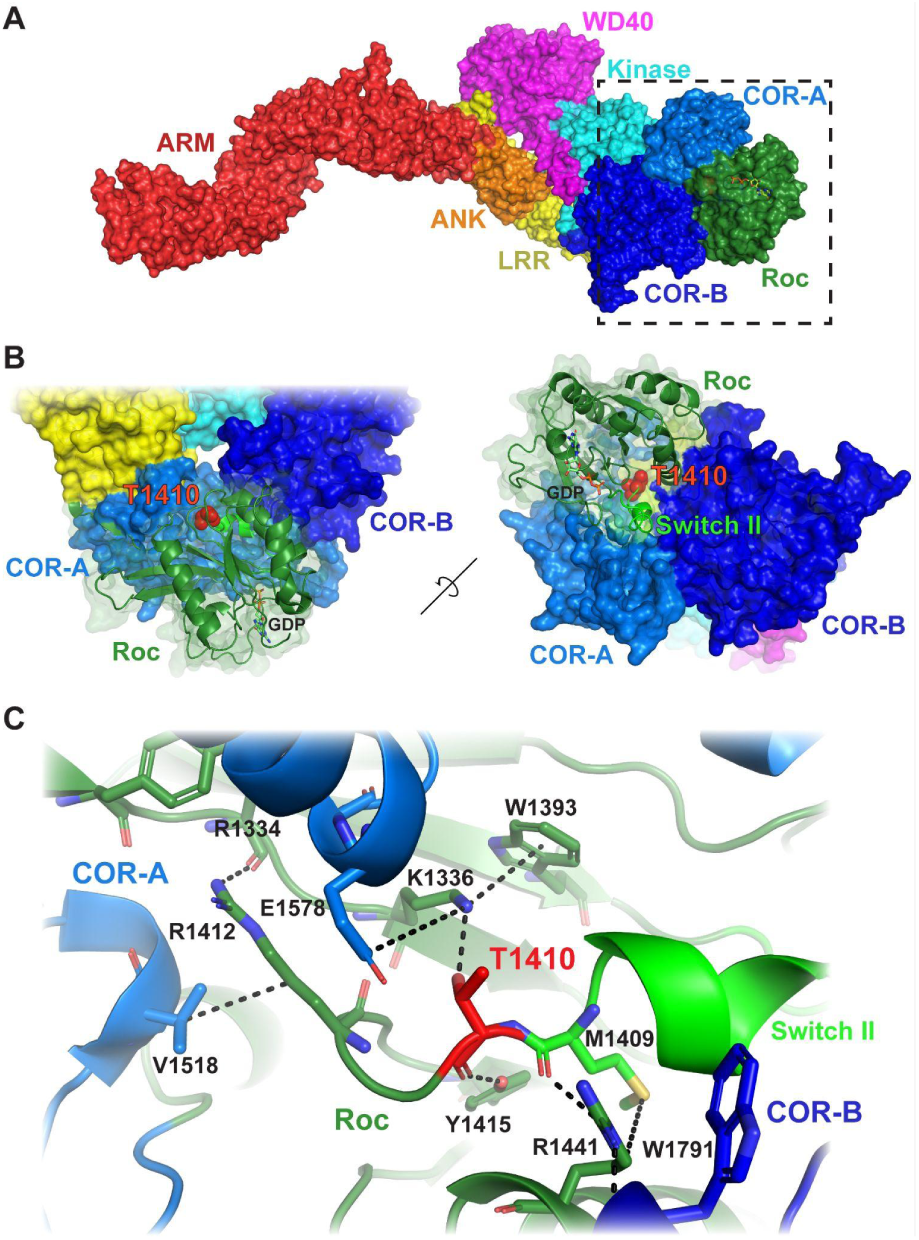
Structural context of Thr1410 within the Roc–COR interface of *LRRK2*. (A) Full-length LRRK2 cryo-EM structure (PDB: 8FO2) showing domain architecture: Roc (dark green), COR-A (blue), COR-B (dark blue), Armadillo (red), Ankyrin (orange), LRR (yellow), kinase (cyan), and WD40 (magenta). (B) Close-up of the Roc/COR region highlighting Thr1410 (red) at the end of the Switch II helix (bright green) and its spatial relationship to both COR lobes. (C) Detailed view of the Thr1410-centred interaction network (based on PDB: 7LI4). Thr1410 hydrogen-bonds with Lys1336 and forms backbone contacts with Tyr1415 and Arg1441; Arg1441 links to Trp1791 in COR-B. Additional stabilising interactions involving Lys1336, Trp1393, Glu1578, Met1409, Arg1412, Arg1334, and Val1518 are shown.

## DISCUSSION

In this study, we present the most comprehensive genome-wide analysis of PD risk in African and African admixed populations to date. By leveraging the individual-level data from GP2 with large-scale summary statistics from collaborative efforts with 23andMe and MVP, we expanded the available cohort size relative to prior work. We identified several genome-wide significant loci across analyses. These findings not only confirm and extend the previously reported *GBA1* signal rs3115534 but also highlight additional loci that emerge only after increasing the representation of individuals with African ancestry in genetic studies of PD.

### *SNCA* as PD GWAS Hit in Global Populations

This is the first time the *SNCA* locus has passed genome-wide significance in the AFR population. The *SNCA* rs356182 variant is a well-established risk factor previously identified in European^6,3^, Latino ^7,8^, and multi-ancestry studies^5^. Its replication here confirms *SNCA* as a trans-ancestry risk locus, fundamental to PD pathology, and importantly, the primary common variant association at the S*NCA* locus appears to be the same signal across populations. *SNCA* encodes alpha-synuclein, a protein central to PD pathogenesis, and point mutations cause early-onset autosomal-dominant PD, while gene overexpression contributes to late-onset and sporadic forms of the disease^51,52^. Misfolding and aggregation of alpha-synuclein disrupt critical cellular processes, including synaptic vesicle function, ultimately leading to neuronal death^53,54^.

### *GBA1* as major risk gene in African ancestry

The most prominent and consistently replicated signal across all analyses was the intronic *GBA1* variant rs3115534. This variant was the single genome-wide significant association in the AAC-only meta-analysis and remained the strongest signal in both the AFR-only and combined AFR/AAC analyses. These results extend our previous work^30^, which first identified rs3115534 as an African-enriched risk allele with effects on PD susceptibility, age at onset, and GCase activity^31^. *GBA1*, with additional lysosomal-related loci identified in this study, reinforces the importance of lysosomal biology in PD risk and, importantly, nominates African ancestry individuals for *GBA1* targeted clinical trials and therapies.

### *SCARB2* as African GWAS Hit Implying Further Lysosomal Risk

The *SCARB2* association (rs11547135) represents the first genome-wide significant signal at this locus in African populations. Follow-up analyses showed that this signal is identical to one of the *SCARB2* signals (**Supplementary Figure 2**). In parallel with our findings, a recent multi-ancestry analysis of GP2 and AMP-PD cohorts, conducted by Sun and colleagues, confirmed the established *SCARB2* GWAS signals rs6812193 and rs6825004 in Europeans and identified rs11547135 as a novel, independent intronic risk variant in GP2’s European cohort, along with ancestry-specific rare-variant burdens and single-variant effects in African American, Ashkenazi Jewish, and East Asian populations^55^. *SCARB2* might contribute to PD risk through a pleiotropic effect of common and rare alleles whose frequencies and effect sizes vary across ancestries. Overall, the genome-wide prioritization of *GBA1*, *SCARB2*, and *SNCA* in these populations points toward a strong lysosomal aspect of risk that may be particularly pronounced in individuals with PD of African populations, suggesting that defects in GCase trafficking and the subsequent failure to clear alpha-synuclein aggregates represent a potential driver of pathogenesis.

### Opening up the Potential for *LRRK2* Trials in African ancestry individuals

When investigating the African-only cohort, we also observed a significant association at *LRRK2* driven by the missense variant rs72546327 (beta = 0.55; p = 1.72E-8), a signal that remained nominally associated in the full AFR/AAC meta-analysis (beta = 0.40; p = 6.51E-7). Mechanistically, *LRRK2* dysfunction is due to perturbation of its kinase and GTPase activities, which are influenced by interactions with cellular membranes, microtubule filaments, and 14-3-3 proteins, as well as by upstream effectors such as Rab29 and VPS35^50^. Previously, across the three ancestry groups (White, Asian, and Arab-Berber), rs72546327 was observed only in individuals of European descent, appearing in 0.07% of cases and 0.02% of controls, consistent with a rare variant of uncertain but potentially increased risk^56^. In gnomAD, rs72546327 is observed primarily in individuals of African/African American ancestry, with an allele frequency of 2.1%, whereas frequencies in all other ancestry groups are below 0.2% or absent (accessed November 2025). *LRRK2* is a gene with well-known ancestral heterogeneity, with population-enriched pathogenic and susceptibility variants. For example, the globally distributed pathogenic mutation p.G2019S resides on a common haplotype of likely Middle Eastern origin, whereas the risk alleles p.G2385R and p.R1628P occur almost exclusively in Asian populations^57^.

Interestingly, the observed effect size of *LRRK2* p.T1410M is more reminiscent of the common coding risk variants *LRRK2* p.G2385R (chr12:40363526:G:A; rs34778348) and *LRRK2* p.R1628P (chr12:40320043:G:C; rs33949390), identified as prominent risk factors in Asian populations^26,49^, than of the stronger disease-linked variants such as *LRRK2* p.G2019S. The observation of three common protein-coding risk variants in *LRRK2* (p.G2385R, p.R1628P, and p.T1410M) raises the question of whether these variants may have conferred a previous selective advantage. Given the proposed role of *LRRK2* in lung function and inflammation, it is plausible that these variants may influence response to infectious disease^58,59^. In addition, given the location of p.T1410M inside the protein structure (**Figure 5**), this variant is of high interest for functional follow-up. The continued investment in *LRRK2*-targeted trials suggests that this hypothesis warrants further investigation.

### Additional novel African ancestry GWAS loci

Our analysis identified additional loci that warrant further investigation. In the AFR-only analysis, we observed an association at rs12302417, located within the non-coding pseudogene *RPL10P13* on chromosome 12. This variant lies approximately 35 Mb from the *LRRK2* locus and unsurprisingly shows no linkage disequilibrium with the *LRRK2*/*SLC2A13* signal in African populations. However, conditional analysis revealed that this signal is not independent of the *LRRK2* risk variant; when conditioning on the *LRRK2* p.T1410M variant (rs72546327), the association at rs12302417 dropped below genome-wide significance (p = 5.516E-08). We also detected a novel association on chromosome 16 at rs113244182 (beta = -0.80, SE = 0.141; p = 1.67E-8). To our knowledge, this locus has not been previously implicated in PD or related neurodegenerative conditions, and the underlying biological mechanism remains unclear. Characterization of this signal will require replication in independent African-ancestry cohorts and functional annotation to identify candidate genes and regulatory elements in the region.

### Limitations

Variants that are rare or absent in European cohorts may reach higher frequencies in African populations and, consequently, provide the statistical power necessary to detect associations that would otherwise remain unseen. As sample sizes grow and sequencing-based approaches become more accessible, these preliminary signals may resolve into validated risk loci with novel biological insights.

Despite these advances, there are limitations of the current work. The recruitment of PD cases from Africa remains concentrated in West Africa, particularly in Nigeria, and therefore does not capture the continent’s extensive genetic diversity. GP2 has several large ongoing efforts throughout Africa to increase diversity and explore other intra-Africa ancestries.

In addition we used microarray-based imputation, which works well for common variants however will miss rare and structural variation. Larger sequencing-based datasets will be needed to characterize the full spectrum of rare and structural variation, which remains difficult to profile accurately.

Lastly, reliance on summary statistics from 23andMe and MVP limits harmonized quality-control procedures, relatedness filtering, and covariate adjustment across cohorts, and the granularity of ancestry prediction. These constraints may introduce heterogeneity in effect size estimates and reduce power to detect ancestry-specific associations. This is particularly visible in the difference in effect sizes between, for example, the *GBA1* variant rs3115534 (**Supplementary Table 2**). Future efforts will prioritize expanded recruitment across Africa, extensive whole-genome sequencing, and multi-omic characterization to further resolve the functional mechanisms underlying the loci identified here.

### Conclusions

In this study, we performed the largest and most comprehensive genome-wide association study of PD risk in AFR and AAC populations to date, leveraging data from GP2, 23andMe, and the MVP. Our expanded cohort increases the necessary statistical power to robustly characterize the genetic architecture of PD in these underrepresented populations. We confirmed *SNCA* (rs356182) as a trans-ancestry risk locus and validated the intronic *GBA1* variant (rs3115534) as the dominant common genetic risk factor in AFR/AAC individuals, solidifying the central role of lysosomal dysfunction in this population. Additionally, we identified the first genome-wide significant association at the *SCARB2* locus (rs11547135), which encodes LIMP-2 and transports GCase to the lysosome. We also provided evidence for the association of the *LRRK2* missense variant p.T1410M (rs72546327), enriched in African ancestry, and identified two novel genome-wide significant loci on chromosome 12 (rs12302417) and chromosome 16 (rs113244182) that likely represent ancestry-specific risk factors requiring functional validation.

This work marks a significant stride toward a more universal genetic model for PD. The collective prioritization of *GBA1*, *SCARB2*, *LRRK2*, and *SNCA* in African ancestry populations reveals strongly related lysosomal signals in African ancestry populations that drive PD pathogenesis and nominates clear, actionable therapeutic targets. Continued expansion of diverse genomic cohorts remains essential to resolve the novel signals identified here and to ensure that precision medicine can be delivered to all PD patients globally.

## Data Availability

Data used in the preparation of this article were obtained from GP2. Specifically we used Tier 2 data from GP2 (release 11: DOI 10.5281/zenodo.17753486). GP2 data can be accessed through AMP PD (https://amp-pd.org). For the MVP dataset, PD summary statistics from the Million Veterans Program (MVP) were downloaded from dbGAP (accession number: phs002453.v1.p1; analysis accession: pha010400.1). Summary statistics from 23andMe were shared under a collaborative agreement submitted at https://research.23andme.com/collaborate/.
All code generated for this article, and the identifiers for all software programs and packages used, are available on GitHub (https://github.com/GP2code/GP2-AFR-AAC-metaGWAS) and were given a persistent identifier via Zenodo (DOI: 10.5281/zenodo.7888140)

## ACKNOWLEDGEMENTS

This project was supported by the Global Parkinson’s Genetics Program (GP2; https://gp2.org). GP2 is funded by the Aligning Science Across Parkinson’s (ASAP) initiative and implemented by The Michael J. Fox Foundation for Parkinson’s Research (MJFF). For a complete list of GP2 members see https://doi.org/10.5281/zenodo.7904831. This research was supported by the Aligning Science Across Parkinson’s Initiative, the Intramural Research Program, National Institute on Aging, National Institutes of Health, Department of Health and Human Services, project ZO1 AG000949, and the Michael J. Fox Foundation for Parkinson’s Research. This work utilized the computational resources of the NIH STRIDES Initiative (https://cloud.nih.gov) through the Other Transaction agreement - Azure: OT2OD032100, Google Cloud Platform: OT2OD027060, Amazon Web Services: OT2OD027852. We would like to thank the research participants, Paul Cannon, and employees of 23andMe Research Institute for making this work possible. We would also like to thank Dario Alessi and his team for their valuable contributions in functionally contextualizing this work.

## DATA AND CODE AVAILABILITY

Data used in the preparation of this article were obtained from GP2. Specifically, we used Tier 2 data from GP2 (release 11; DOI 10.5281/zenodo.17753486). GP2 data can be accessed through AMP PD (https://amp-pd.org). For the MVP dataset, PD summary statistics from the Million Veteran’s Program (MVP) were downloaded from dbGAP (accession number: phs002453.v1.p1; analysis accession: pha010400.1). Summary statistics from 23andMe were shared under a collaborative agreement, submitted at https://research.23andme.com/collaborate/.

All code generated for this article, and the identifiers for all software programs and packages used, are available on GitHub (https://github.com/GP2code/GP2-AFR-AAC-metaGWAS) and were given a persistent identifier via Zenodo (DOI: 10.5281/zenodo.7888140)

## CONTRIBUTIONS

Mary B. Makarious, Hampton L. Leonard, and Lara M. Lange designed the analysis, analyzed the data, and drafted the initial manuscript.

Kristin Levine, Hampton L. Leonard, Mary B. Makarious, Dan Vitale, Mat Koretsky, and Nicole Kuznetsov contributed to quality control of GP2 genetic data and making it available to researchers.

Njideka U Okubadejo, Huw R. Morris, Cornelis Blauwendraat and Andrew B. Singleton led study design, data logistics, and funding of the study.

Huw Morris, Manuela Tan, Hirotaka Iwaki, Simona Jasaityte, Ellie Stafford, Lietsel Jones, Shannon Ballard, and Claire Wegel (Complex and Compliance Working Groups) designed the GP2 complex study, clinical and genetic data generation and preparation, and legal / data sharing logistics.

All other members of GP2 (contributors) contributed data and made critical revisions to this article.

---

**Supplementary Figure 1.**
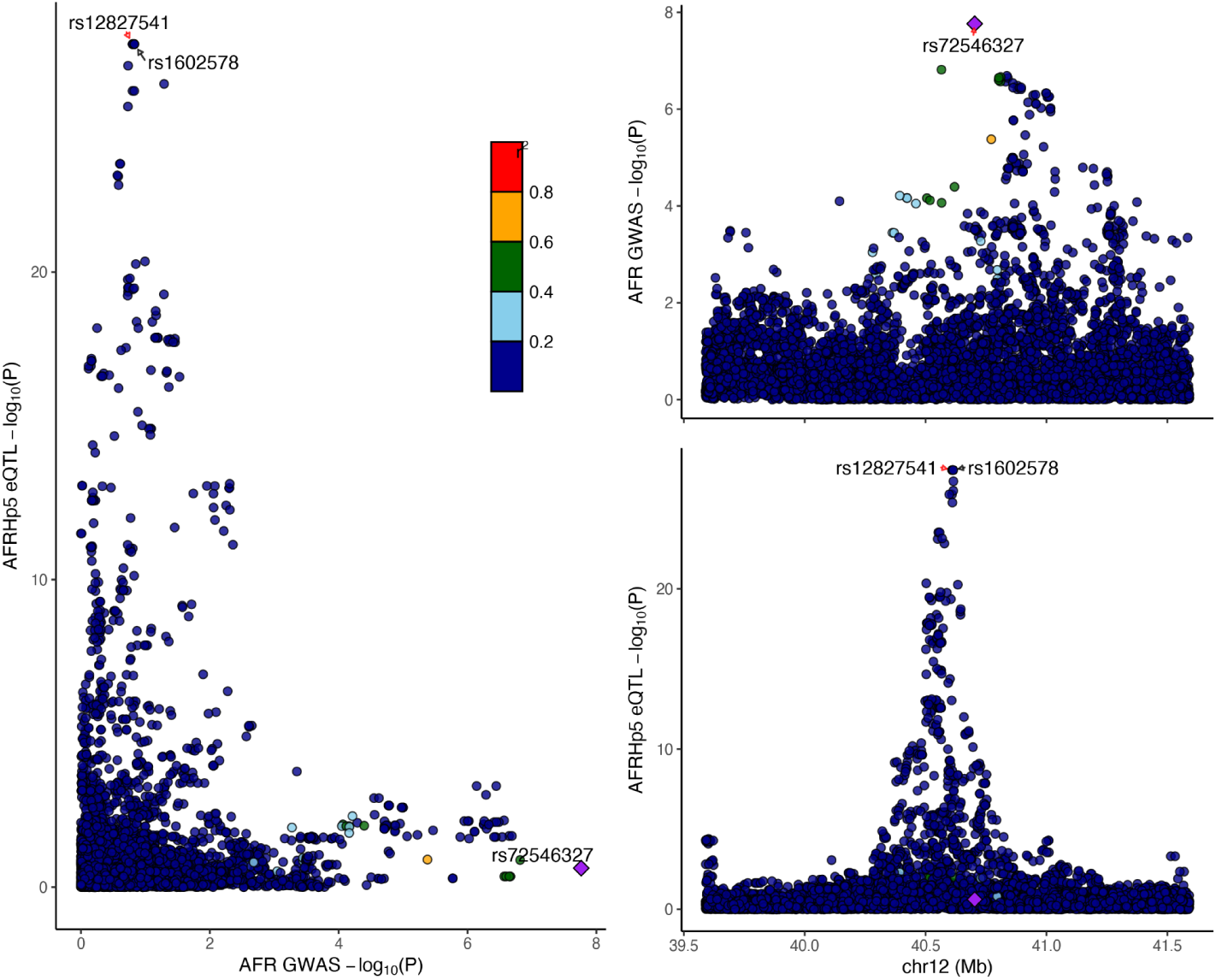
LocusCompare in *LRRK2* region. LocusZoom plot showing African GWAS summary statistics compared against AFRHp5 (>50% African ancestry) whole-blood cis-eQTL summary statistics from Kachuri et al. (2023)

**Supplementary Figure 2.**
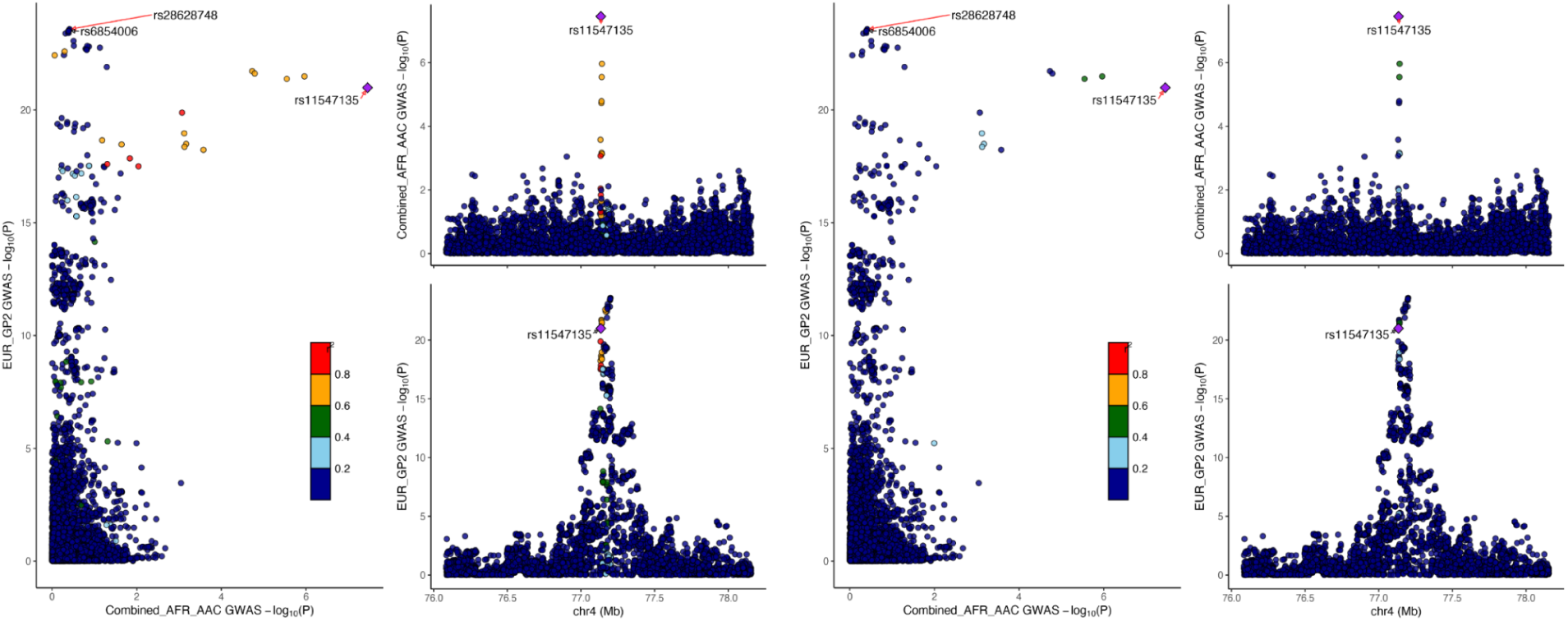
LocusCompare in LRRK2/*SCARB2* region. +/- 1Mb around *SCARB2* GP2 EUR vs Combined AFR/AAC meta-analysis (EUR LD left; AFR LD right)

**Supplementary Figure 3.**
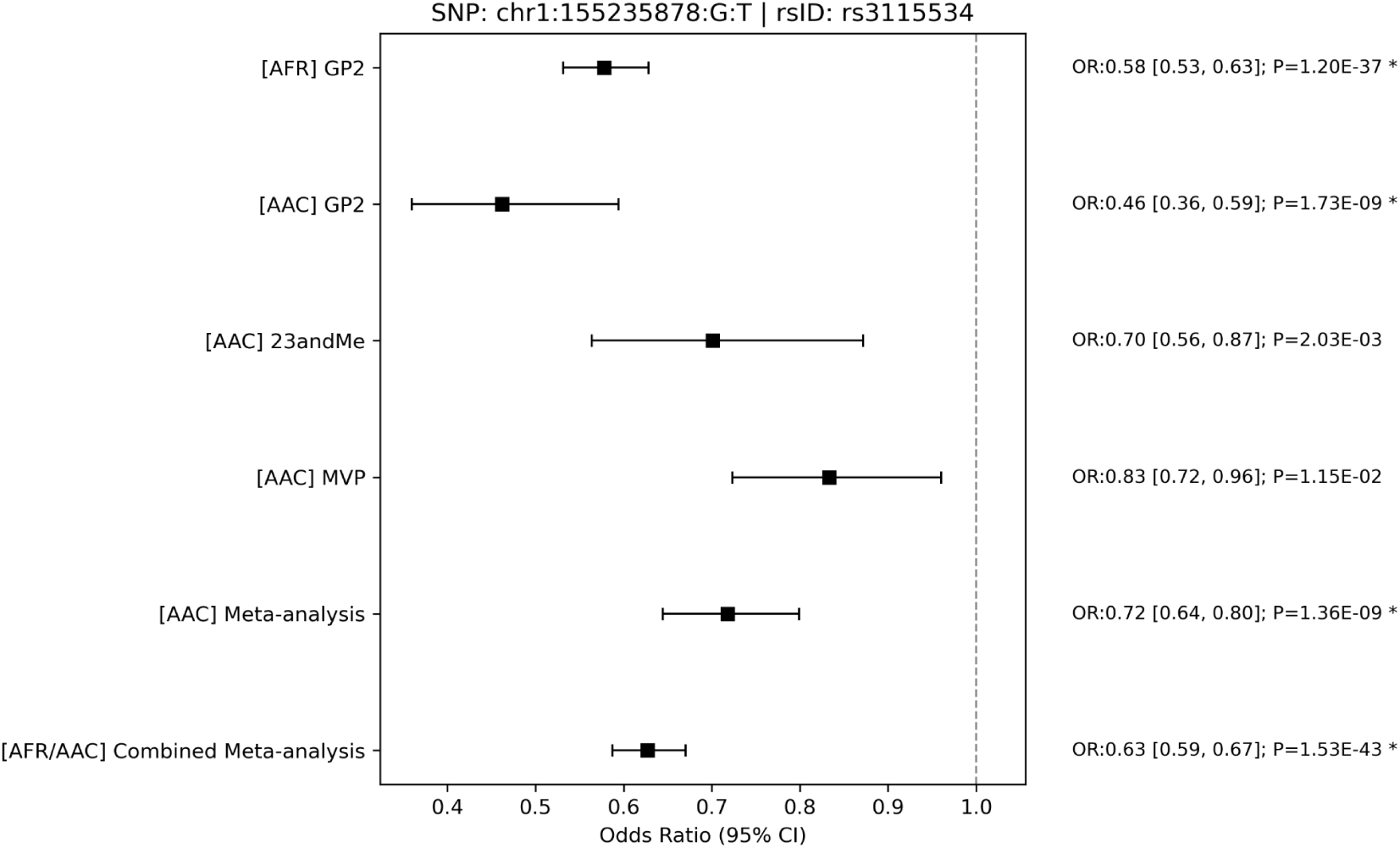
Forest Plot for rs3115534 Across Cohorts. GP2: Global Parkinson’s Genetics Program; MVP: Million Veteran Program; AAC: African admixed ancestry; AFR: African ancestry; OR: Odds Ratio

**Supplementary Figure 4.**
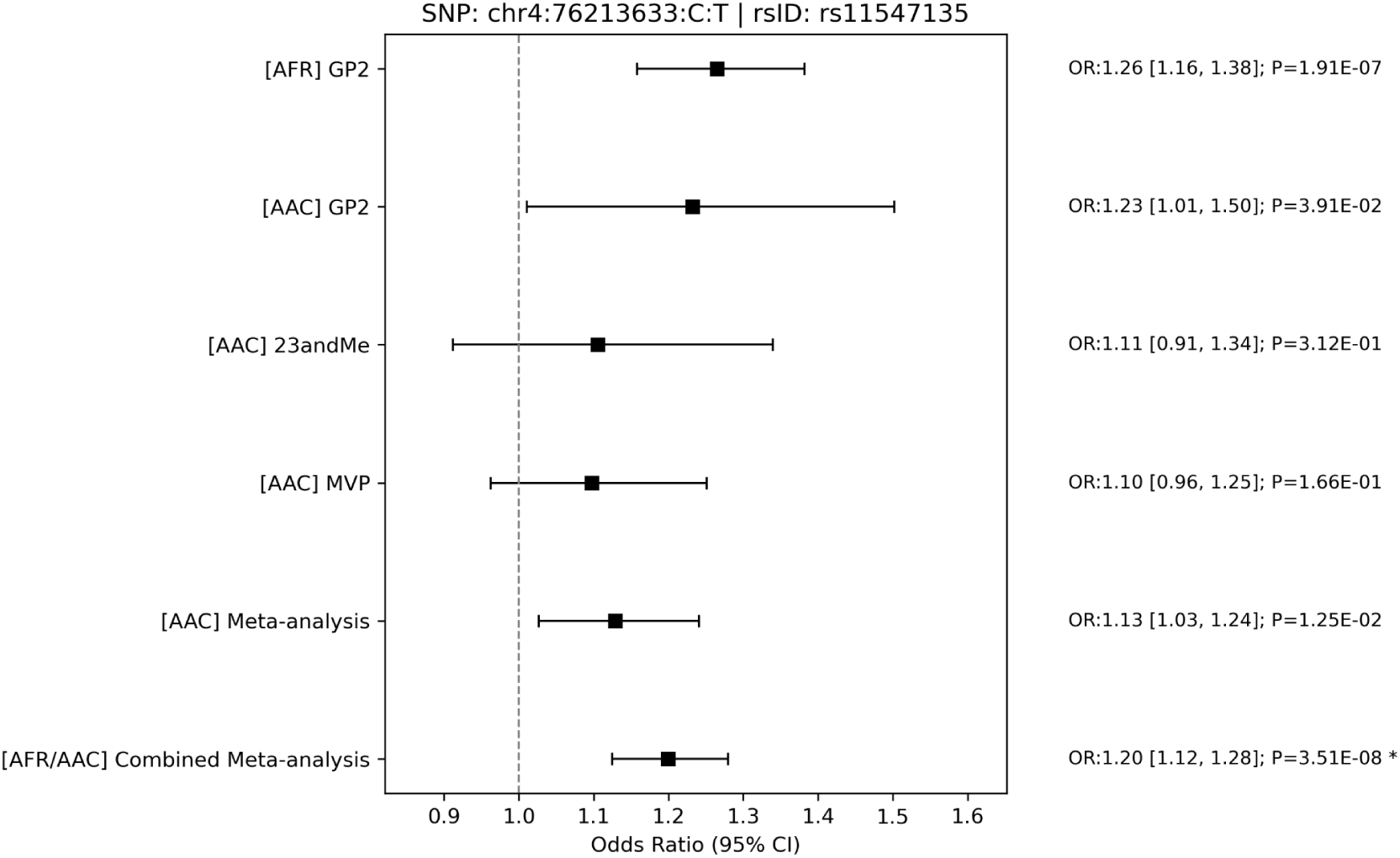
Forest Plot for rs11547135 Across Cohorts. GP2: Global Parkinson’s Genetics Program; MVP: Million Veteran Program; AAC: African admixed ancestry; AFR: African ancestry; OR: Odds Ratio

**Supplementary Figure 5.**
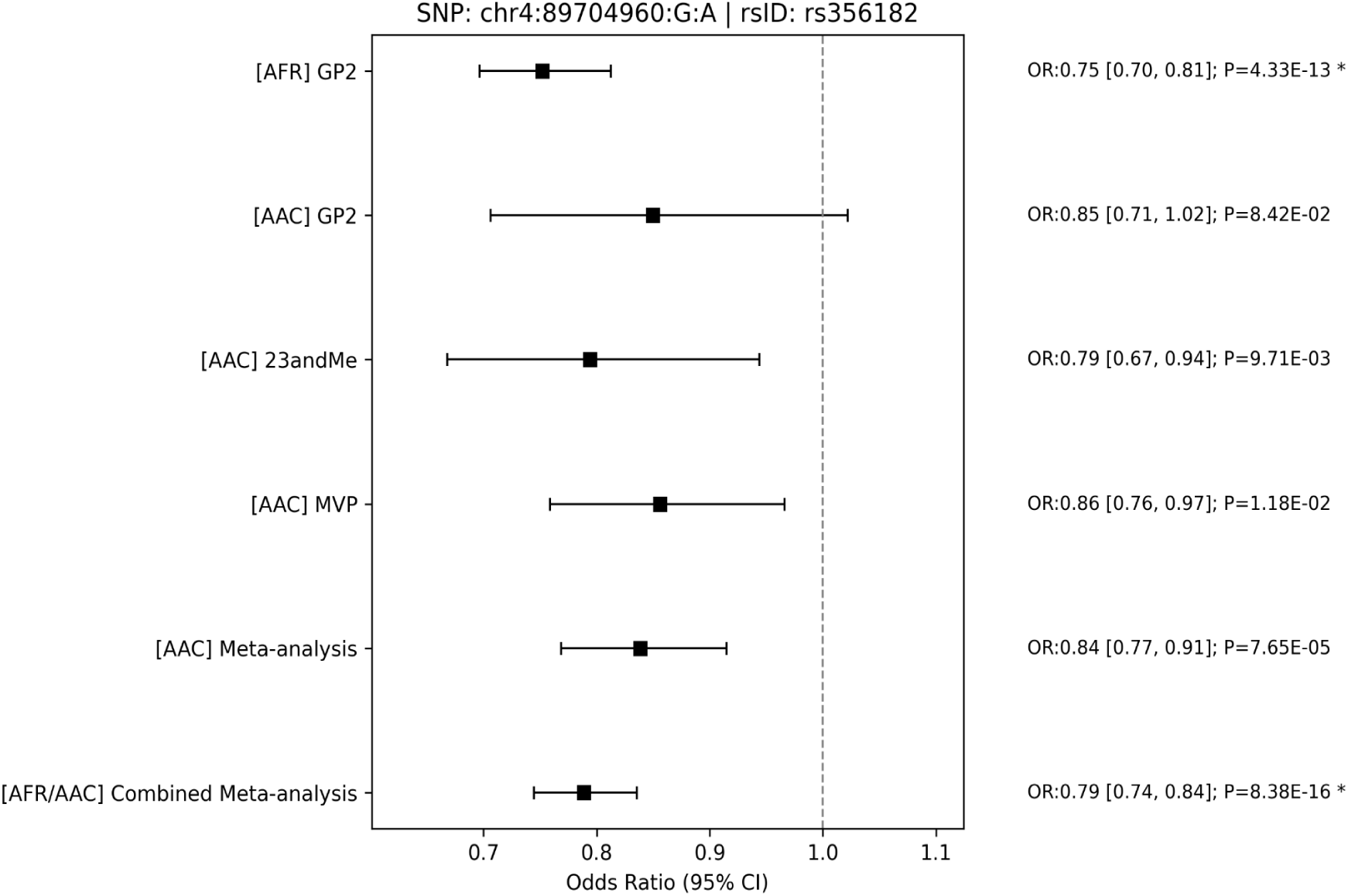
Forest Plot for rs356182 Across Cohorts. GP2: Global Parkinson’s Genetics Program; MVP: Million Veteran Program; AAC: African admixed ancestry; AFR: African ancestry; OR: Odds Ratio

**Supplementary Figure 6.**
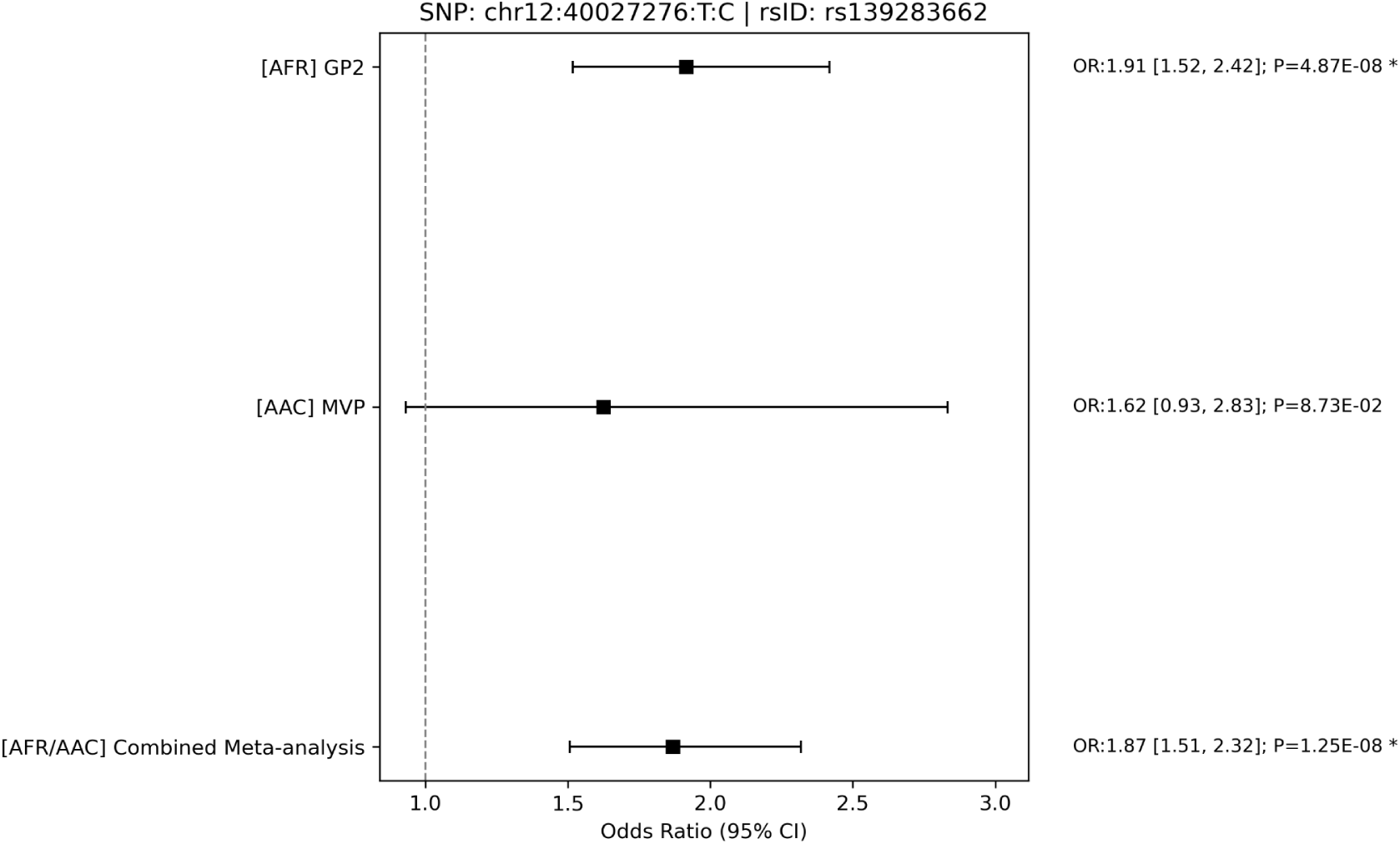
Forest Plot for rs139283662 Across Cohorts. GP2: Global Parkinson’s Genetics Program; MVP: Million Veteran Program; AAC: African admixed ancestry; AFR: African ancestry; OR: Odds Ratio

**Supplementary Figure 7.**
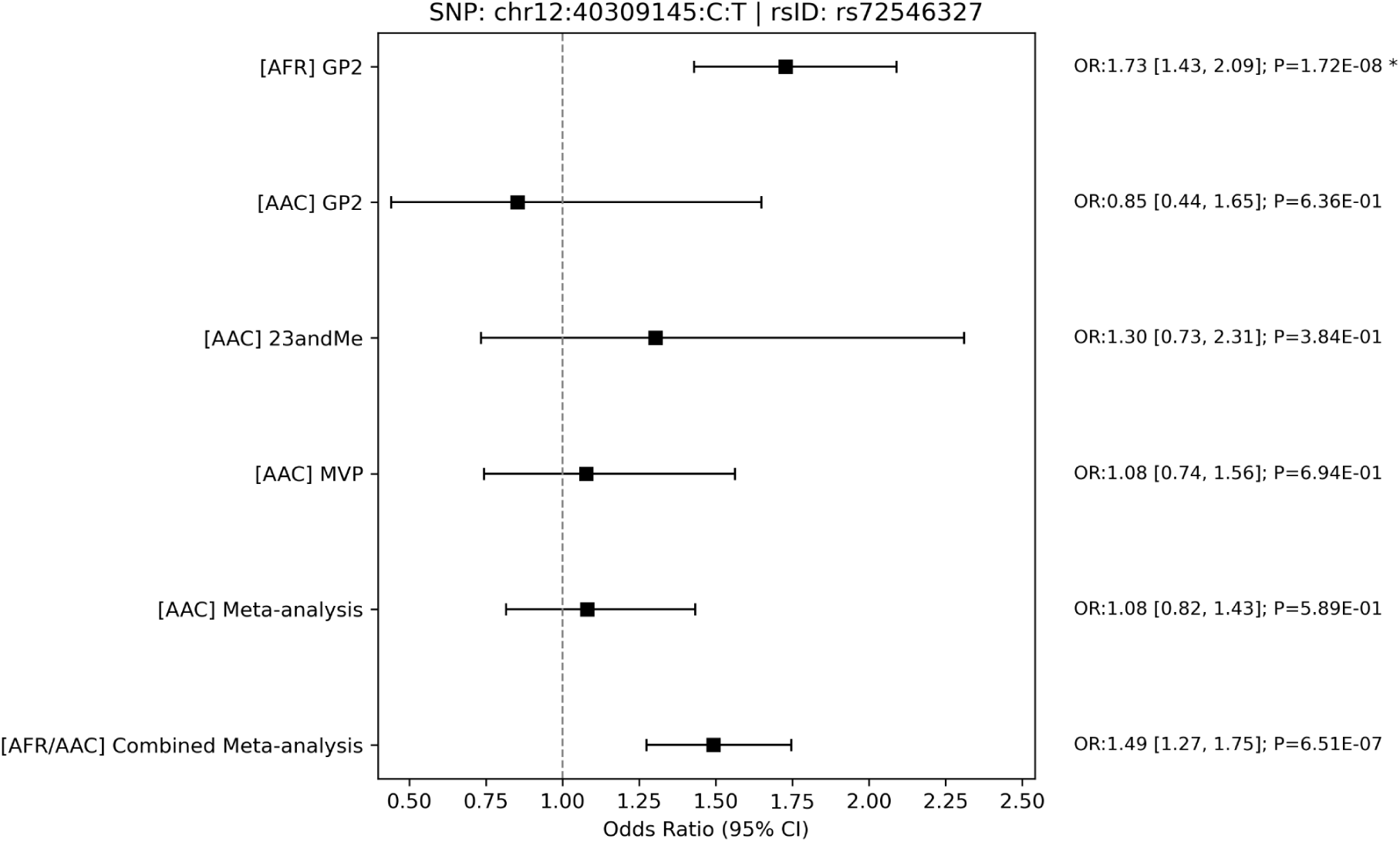
Forest Plot for rs72546327 Across Cohorts. GP2: Global Parkinson’s Genetics Program; MVP: Million Veteran Program; AAC: African admixed ancestry; AFR: African ancestry; OR: Odds Ratio

**Supplementary Figure 8.**
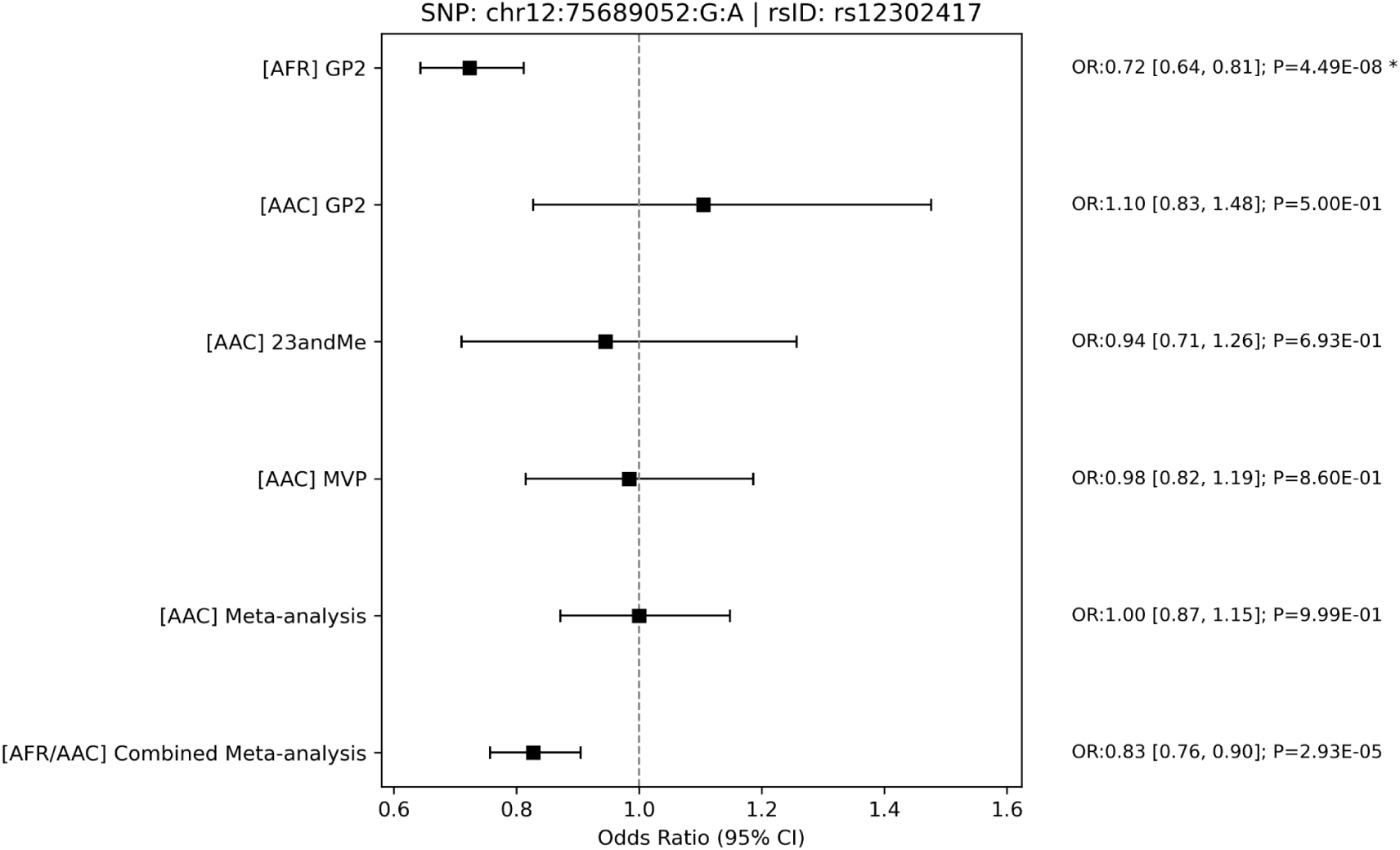
Forest Plot for rs12302417 Across Cohorts. GP2: Global Parkinson’s Genetics Program; MVP: Million Veteran Program; AAC: African admixed ancestry; AFR: African ancestry; OR: Odds Ratio

**Supplementary Figure 9.**
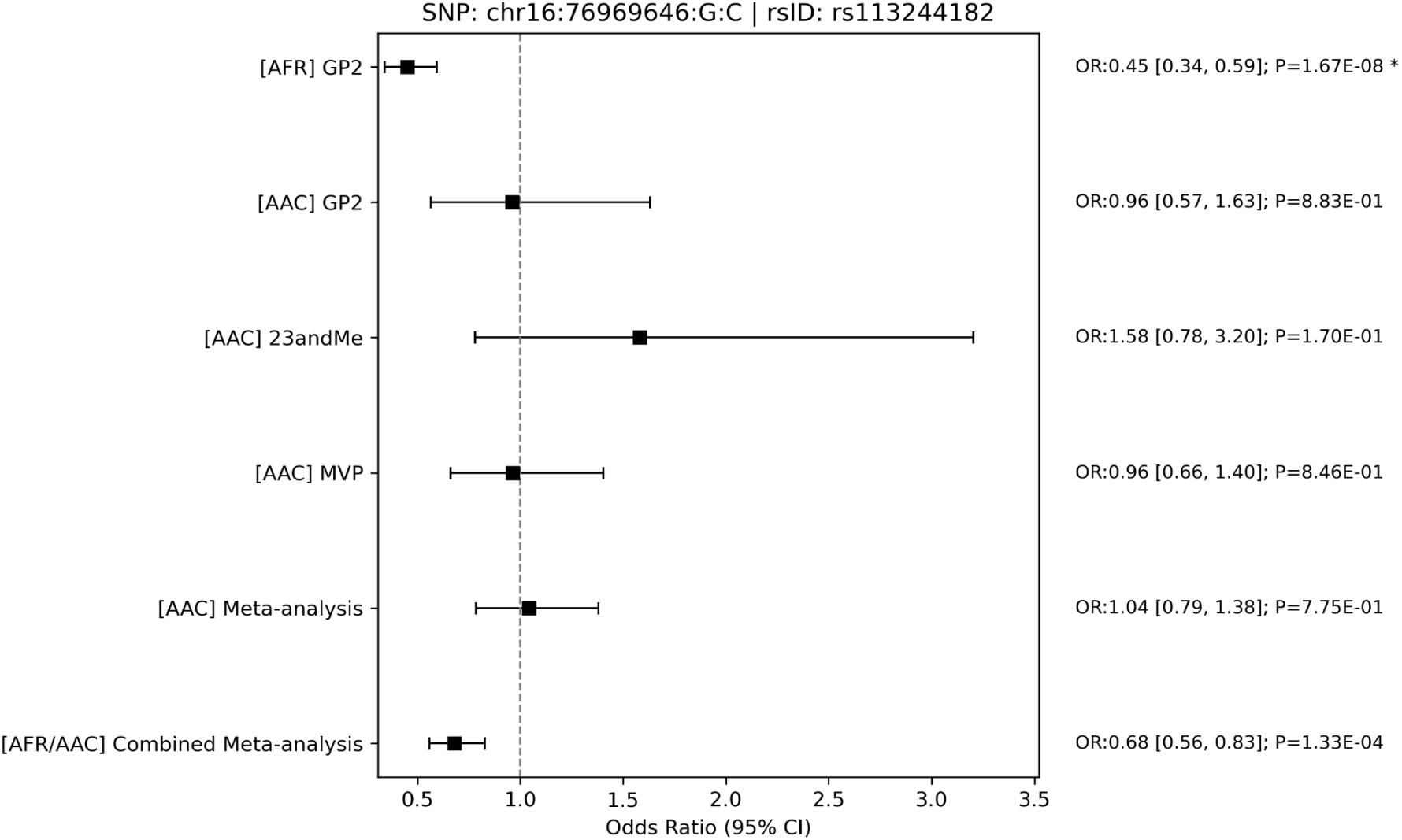
Forest Plot for rs113244182 Across Cohorts. GP2: Global Parkinson’s Genetics Program; MVP: Million Veteran Program; AAC: African admixed ancestry; AFR: African ancestry; OR: Odds Ratio

**Supplementary Figure 10.**
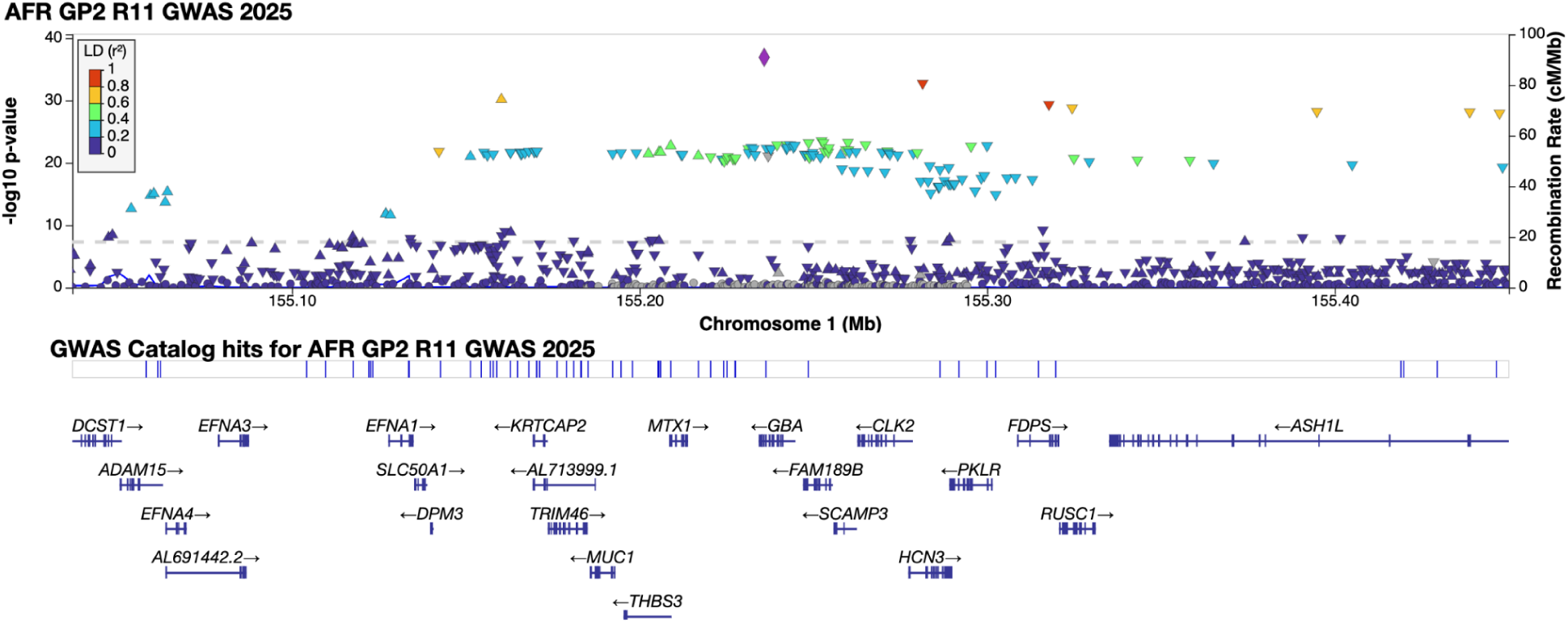
AFR LocusZoom Plot for AFR rs3115534 (chr1:155235878:G:T)

**Supplementary Figure 11.**
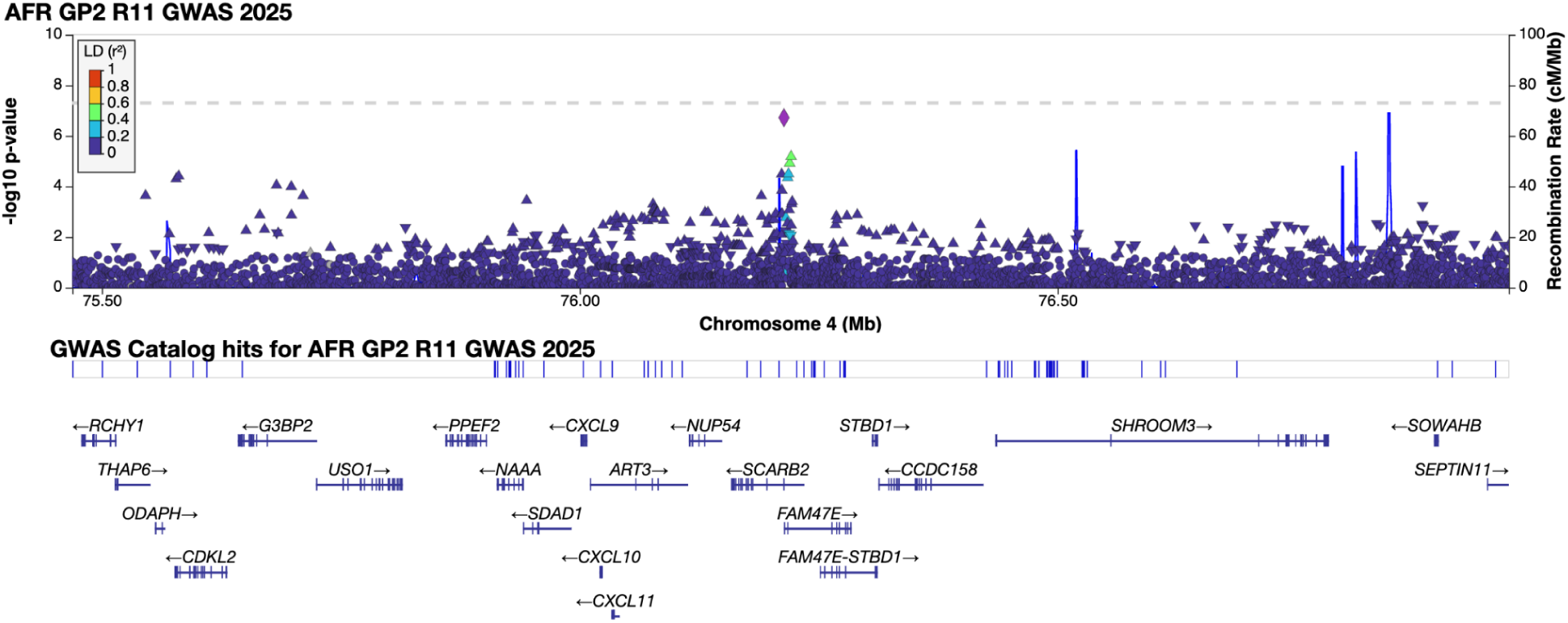
AFR LocusZoom Plot for rs356182 (chr4:89704960:G:A)

**Supplementary Figure 12.**
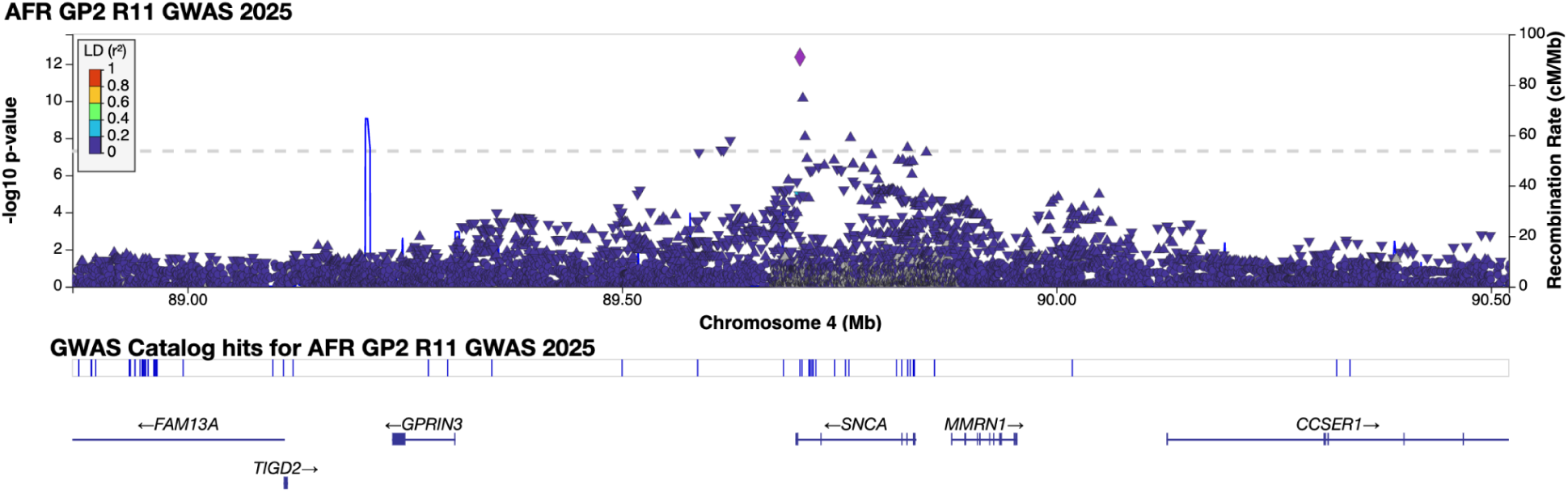
AFR LocusZoom Plot for rs356182 (chr4:89704960:G:A)

**Supplementary Figure 13.**
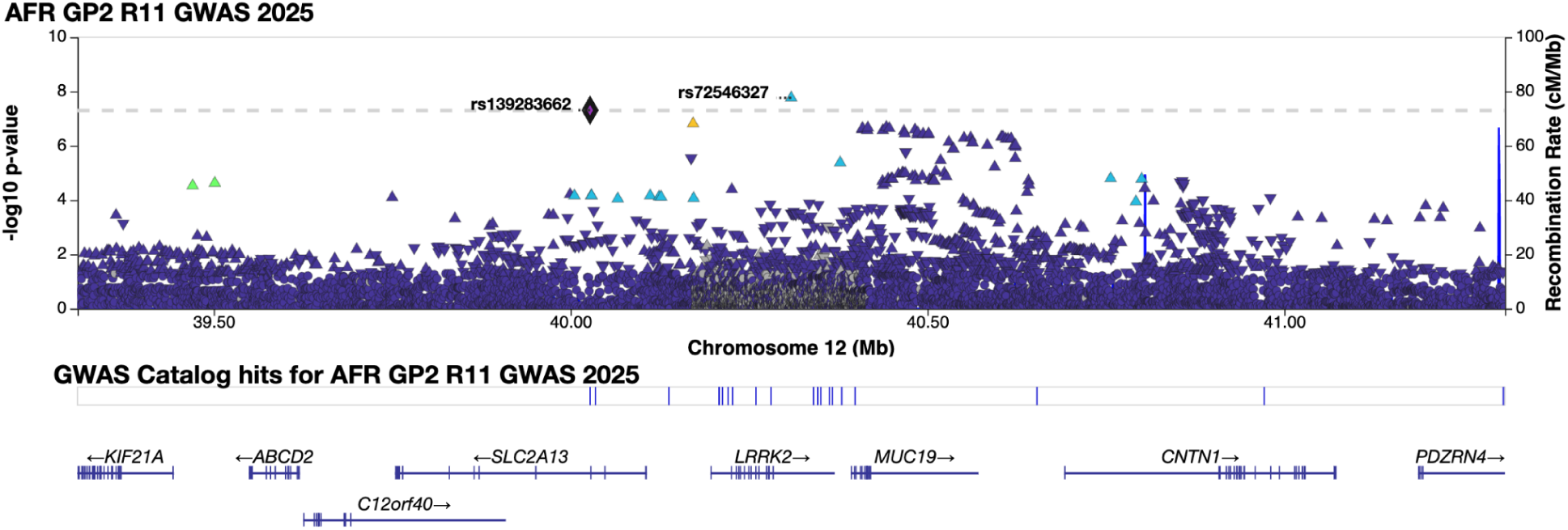
AFR LocusZoom Plot for *LRRK2* locus. rs72546327 (chr12:40309145:C:T) and rs139283662 (chr12:40027276:T:C)

**Supplementary Figure 14.**
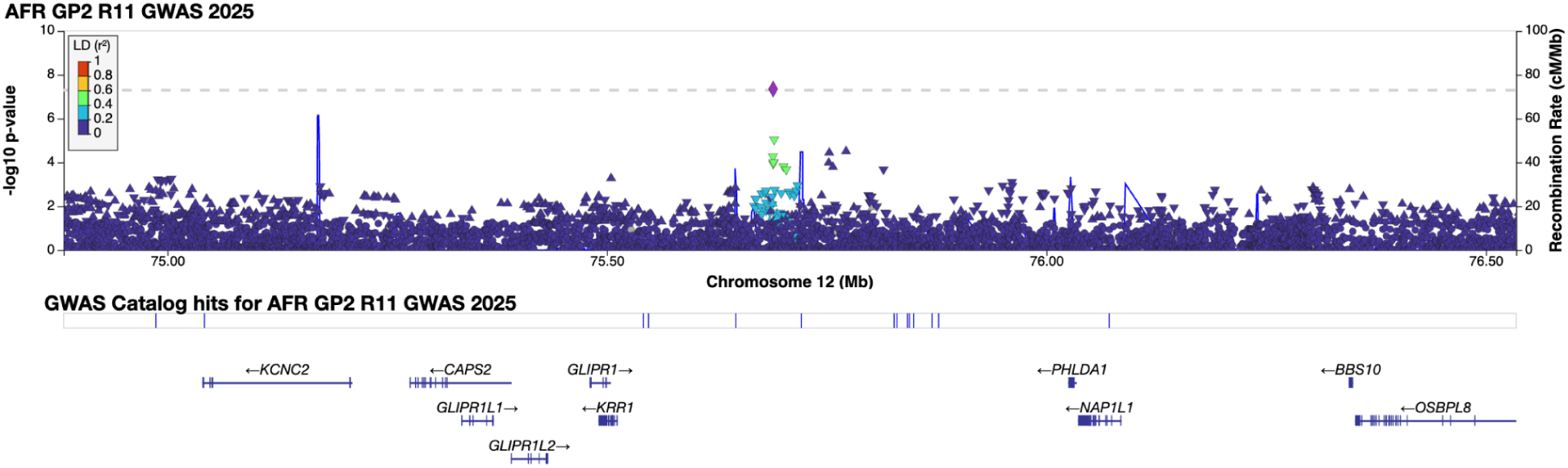
AFR LocusZoom Plot for rs12302417 (chr12:75689052:G:A)

**Supplementary Figure 15.**
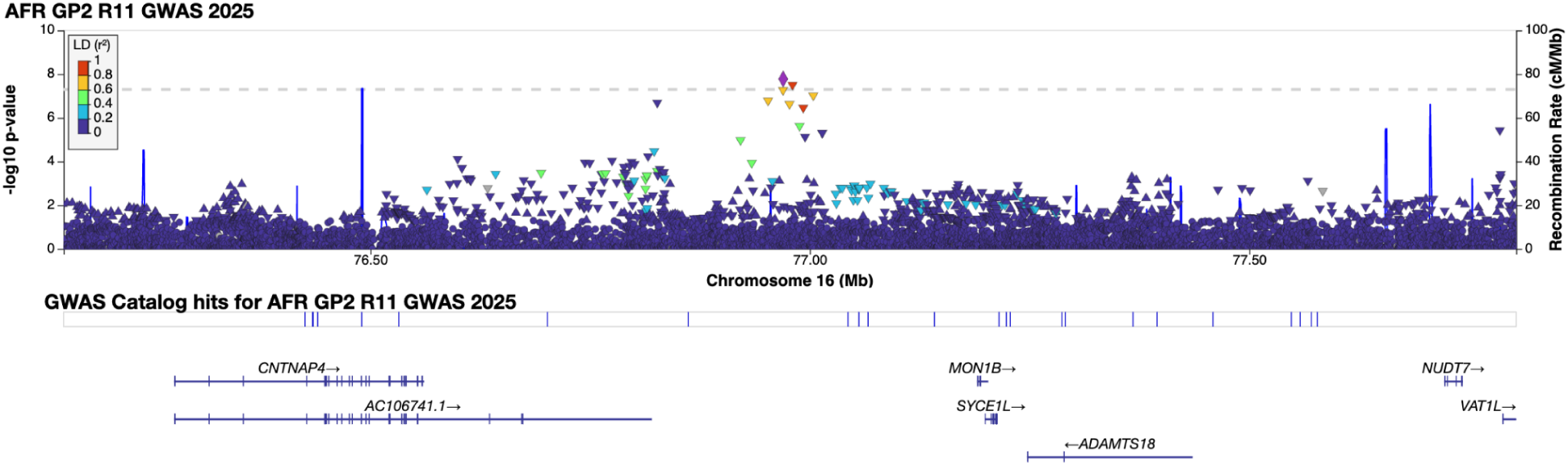
AFR LocusZoom Plot for rs113244182 (chr16:76969646:G:C)

**Supplementary Figure 16.**
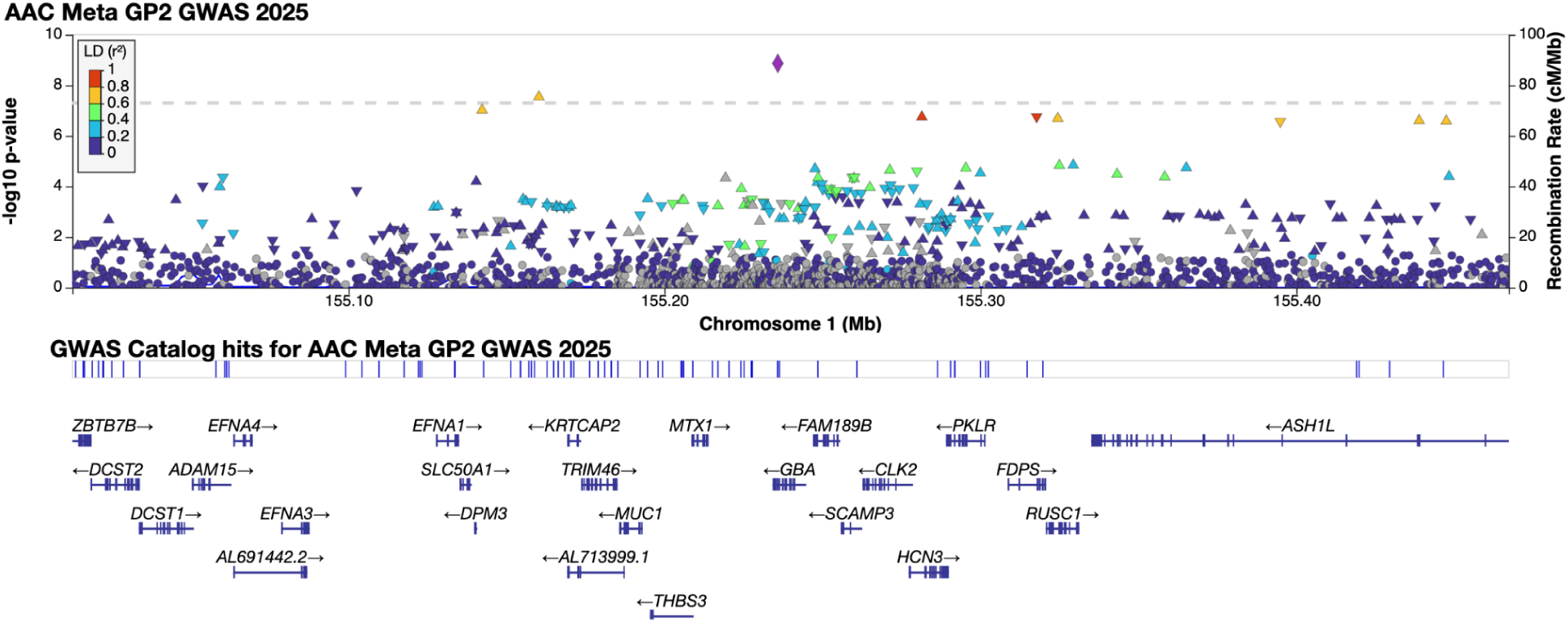
AAC LocusZoom Plot for rs3115534 (chr1:155235878:G:T)

**Supplementary Figure 17.**
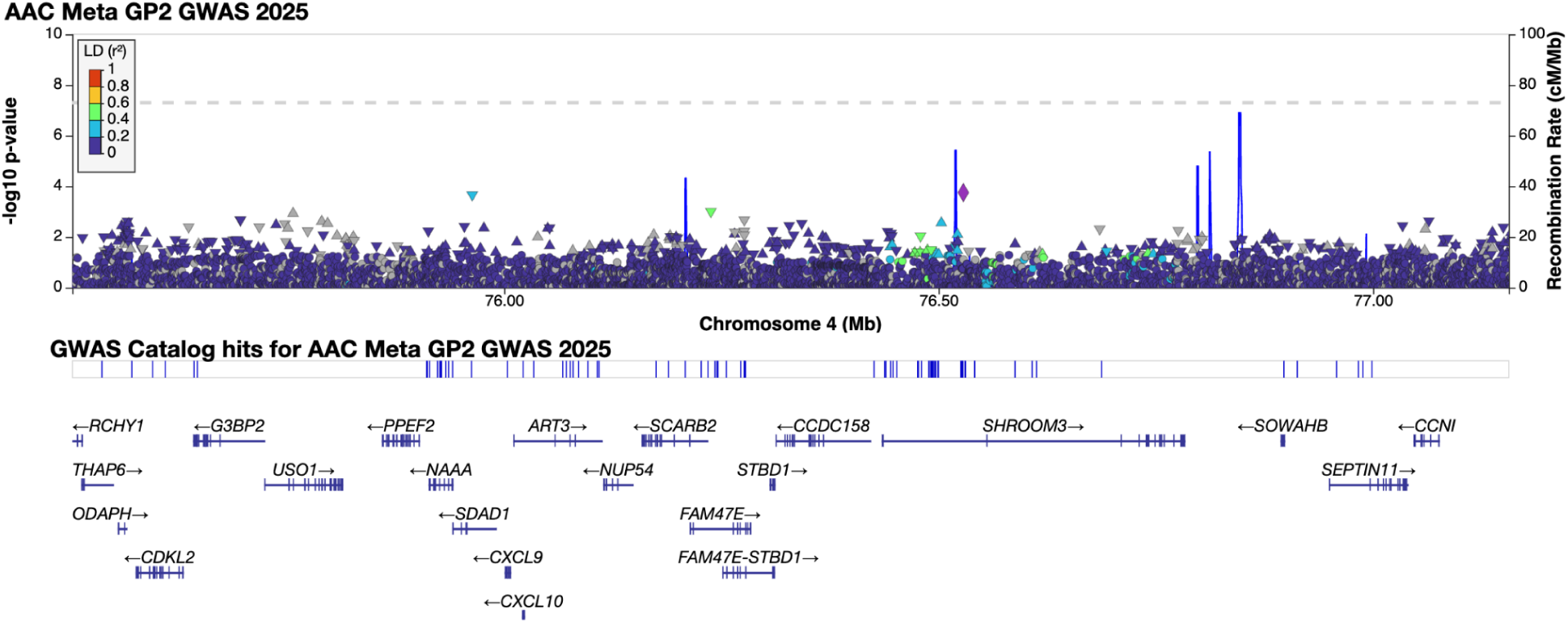
AAC LocusZoom Plot for rs11547135 (chr4:76213633:C:T)

**Supplementary Figure 18.**
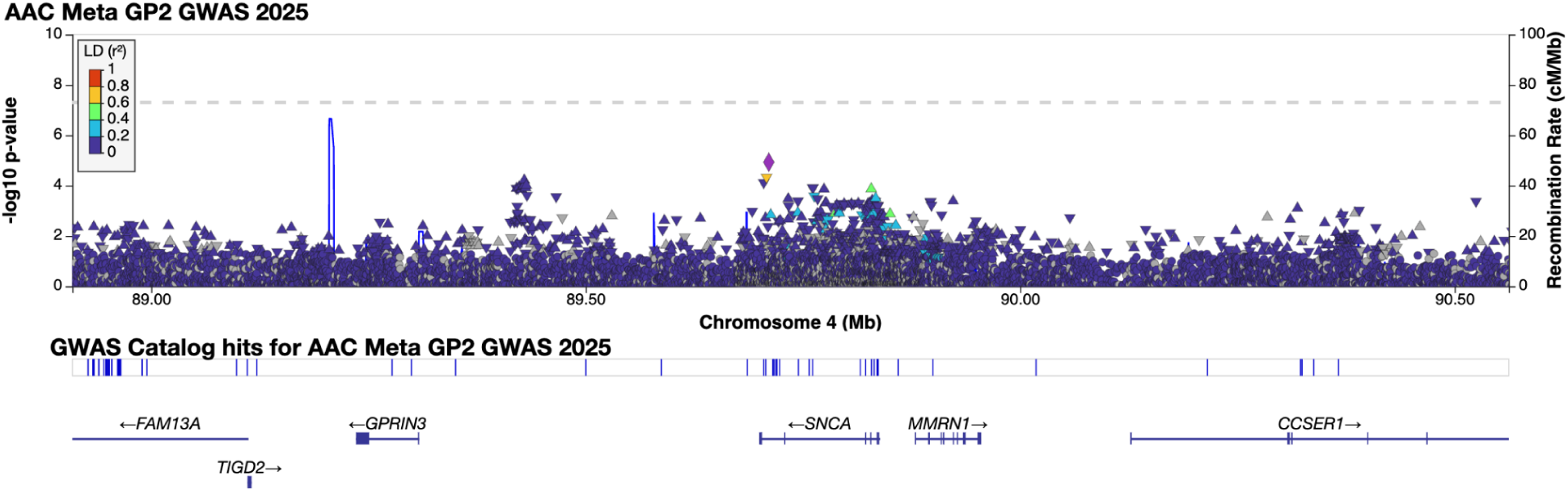
AAC LocusZoom Plot for rs356182 (chr4:89704960:G:A)

**Supplementary Figure 19.**
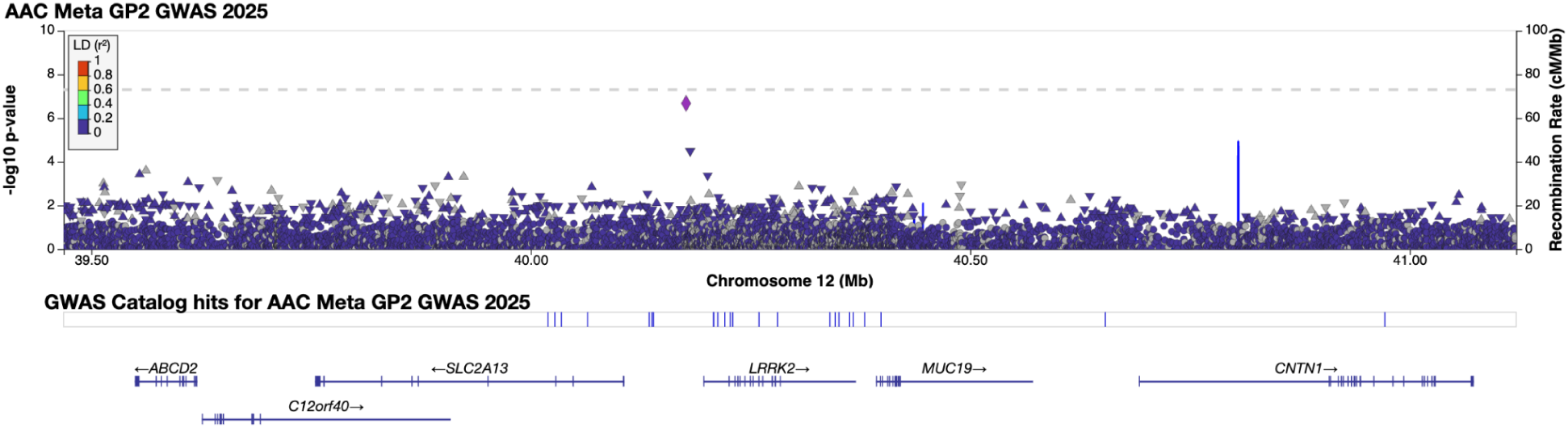
AAC LocusZoom Plot for *LRRK2* locus. rs72546327 (chr12:40309145:C:T) and rs139283662 (chr12:40027276:T:C)

**Supplementary Figure 20.**
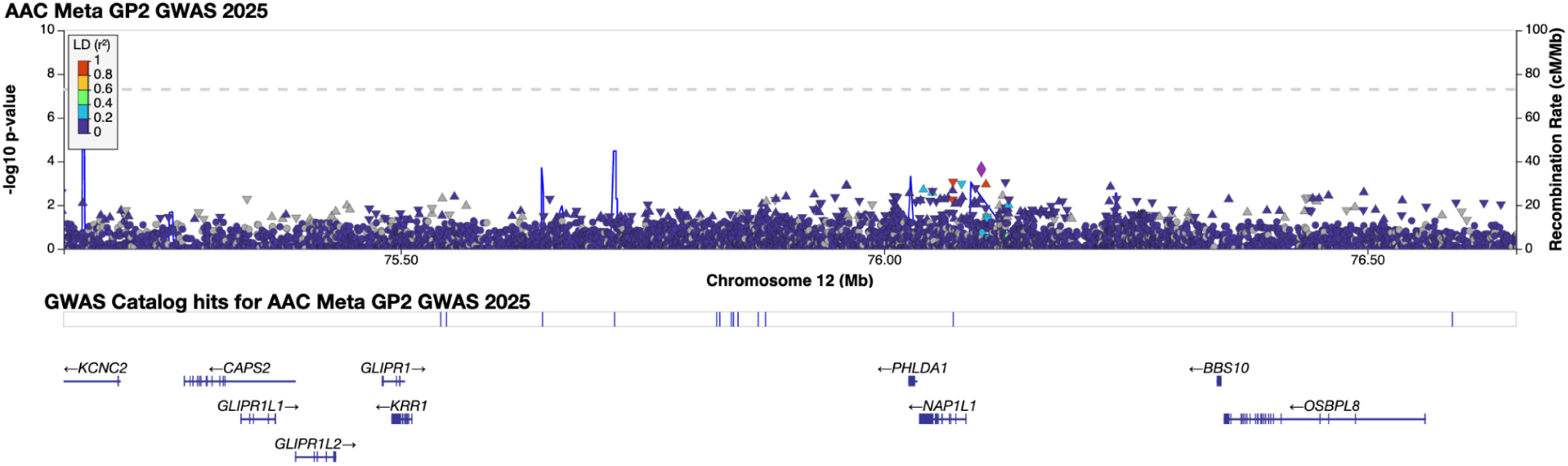
AAC LocusZoom Plot for rs12302417 (chr12:75689052:G:A)

**Supplementary Figure 21.**
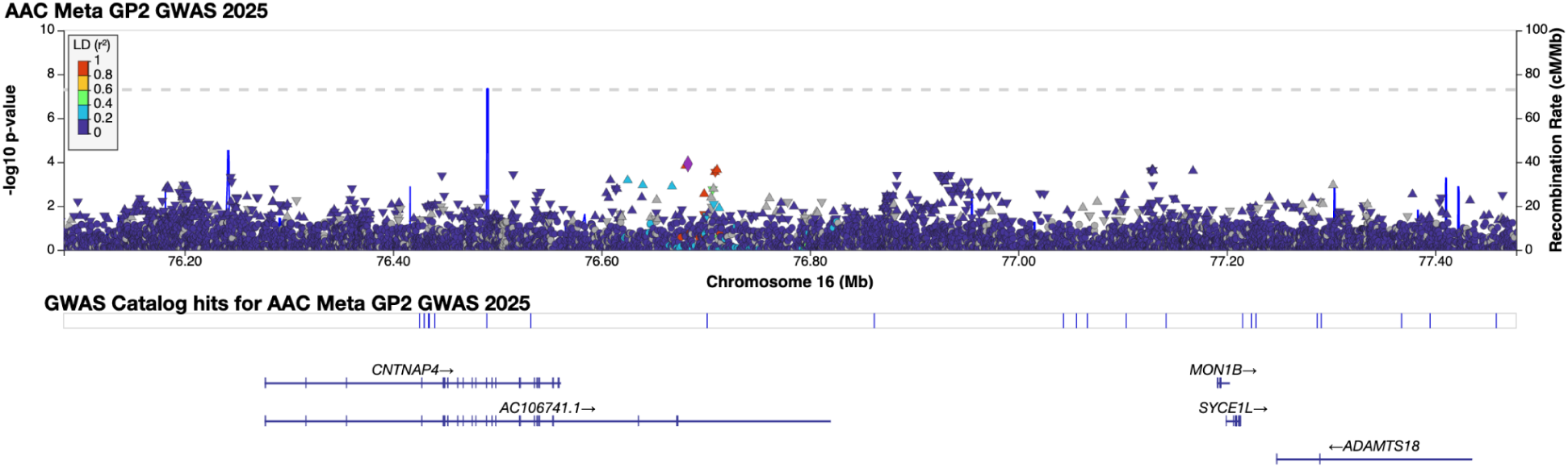
AAC LocusZoom Plot for rs113244182 (chr16:76969646:G:C)

**Supplementary Figure 22.**
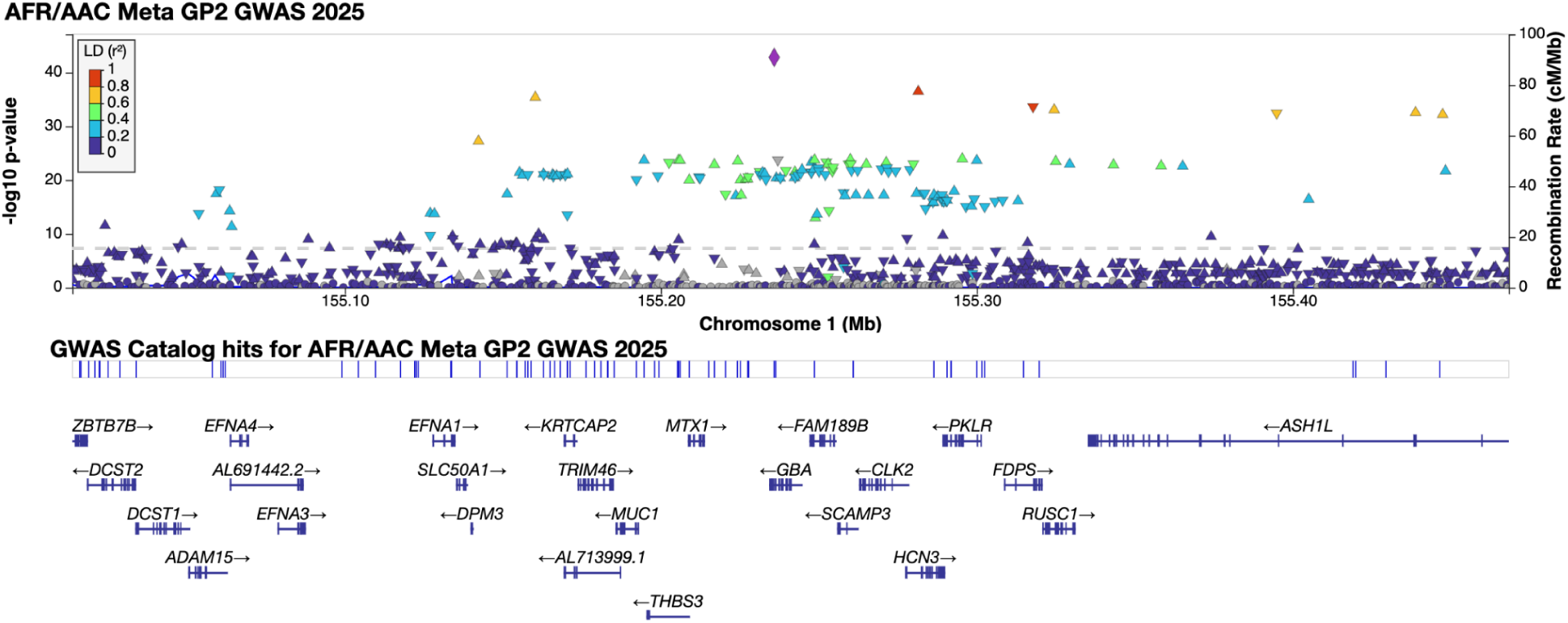
AFR/AAC LocusZoom Plot for rs3115534 (chr1:155235878:G:T)

**Supplementary Figure 23.**
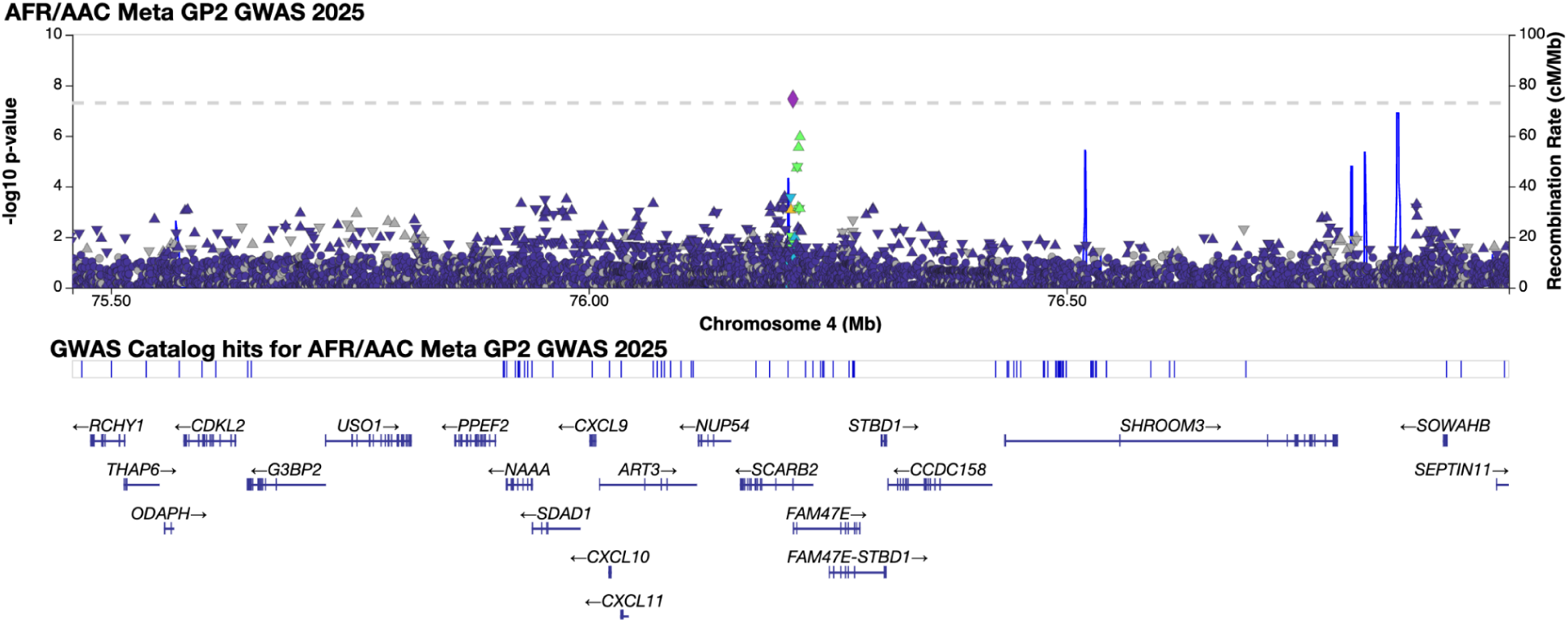
AFR/AAC LocusZoom Plot for rs11547135 (chr4:76213633:C:T)

**Supplementary Figure 24.**
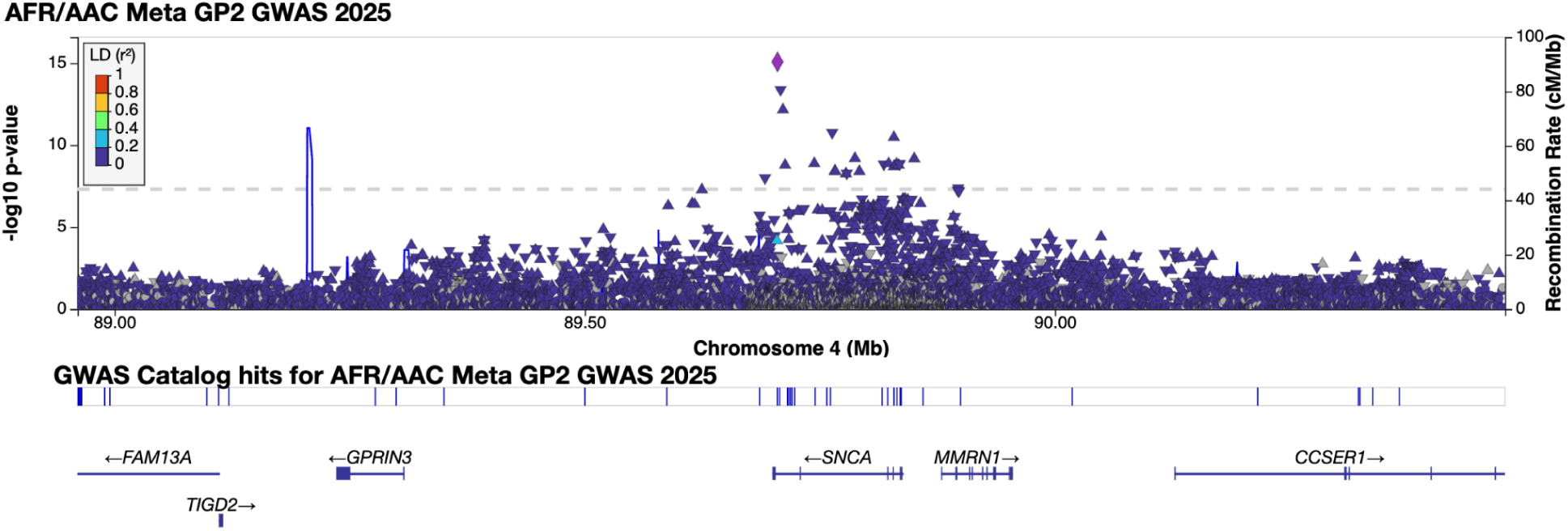
AFR/AAC LocusZoom Plot for rs356182 (chr4:89704960:G:A)

**Supplementary Figure 25.**
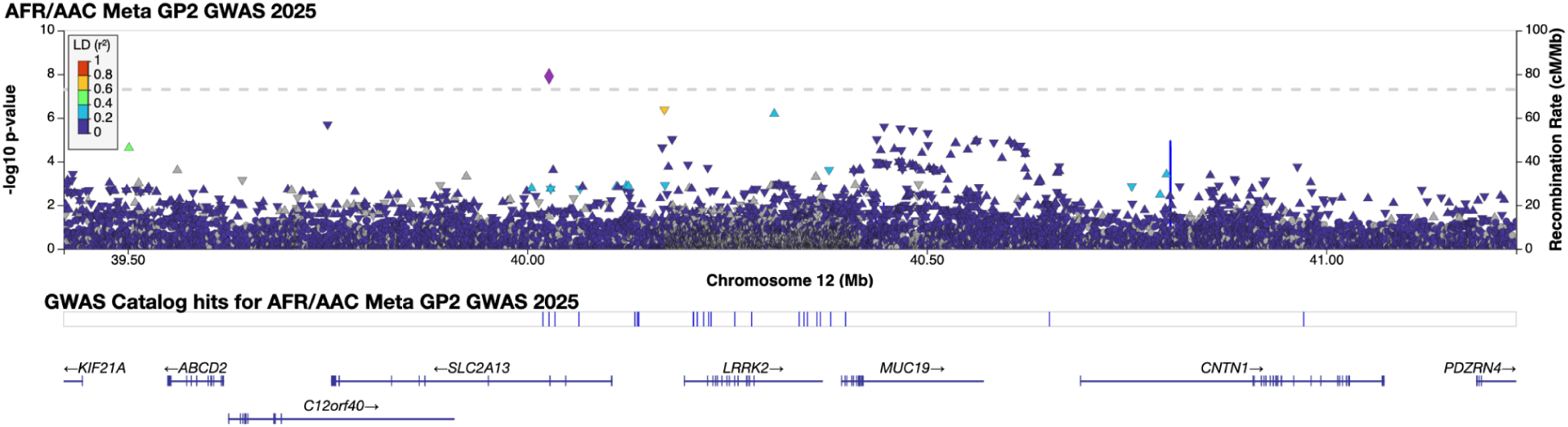
AFR/AAC LocusZoom Plot for *LRRK2* locus. rs72546327 (chr12:40309145:C:T) and rs139283662 (chr12:40027276:T:C)

**Supplementary Figure 26.**
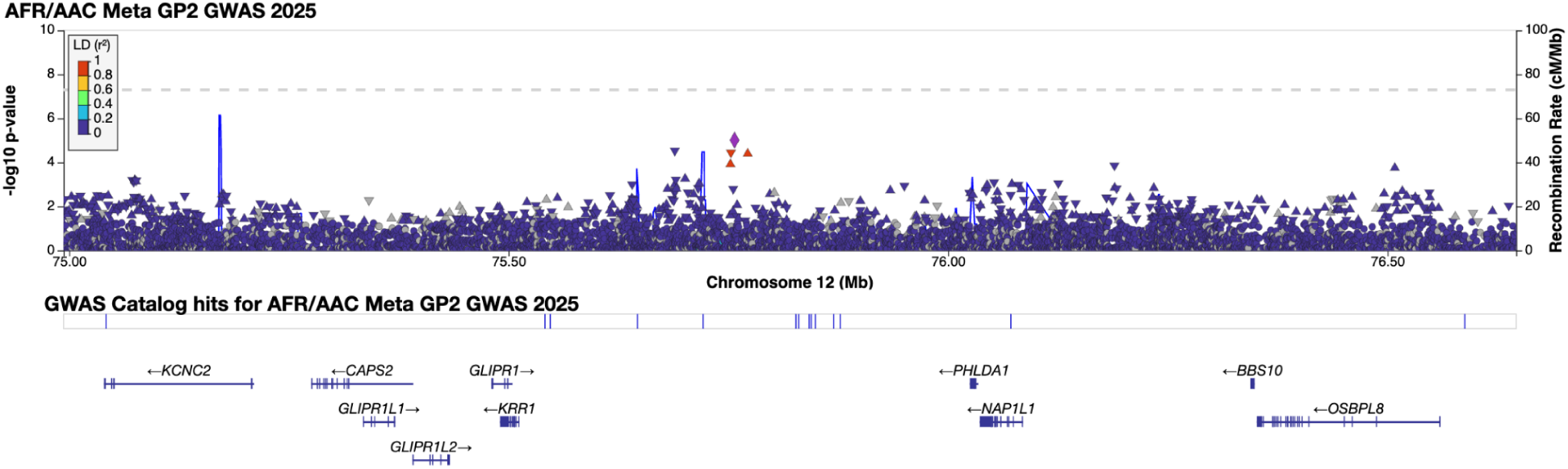
AFR/AAC LocusZoom Plot for rs12302417 (chr12:75689052:G:A)

**Supplementary Figure 27.**
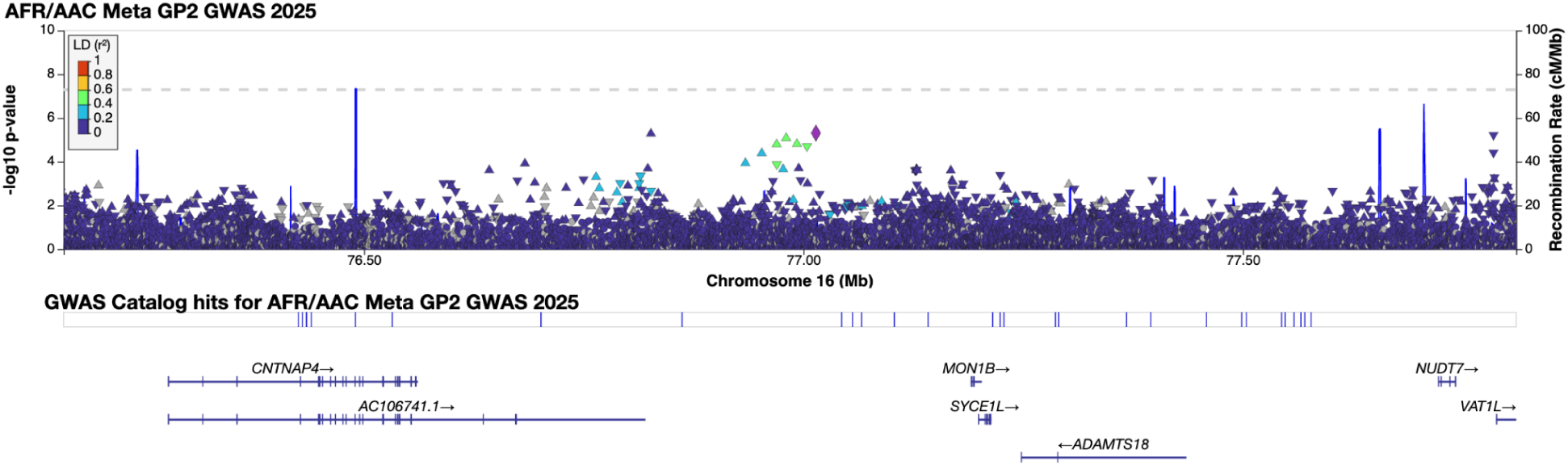
AFR/AAC LocusZoom Plot for rs113244182 (chr16:76969646:G:C)

## REFERENCES

1 GBD 2021 Nervous System Disorders Collaborators. Global, regional, and national burden of disorders affecting the nervous system, 1990-2021: a systematic analysis for the Global Burden of Disease Study 2021. Lancet Neurol 2024; 23: 344–81.

2 Dorsey ER, Sherer T, Okun MS, Bloem BR. The Emerging Evidence of the Parkinson Pandemic. J Parkinsons Dis 2018; 8: S3–8.

3 Leonard HL, Global Parkinson’s Genetics Program (GP2). Novel Parkinson’s Disease Genetic Risk Factors Within and Across European Populations. medRxiv. 2025; published online March 17. DOI:10.1101/2025.03.14.24319455.

4 Foo JN, Chew EGY, Chung SJ, et al. Identification of Risk Loci for Parkinson Disease in Asians and Comparison of Risk Between Asians and Europeans: A Genome-Wide Association Study. JAMA Neurol 2020; 77: 746–54.

5 Kim JJ, Vitale D, Otani DV, et al. Multi-ancestry genome-wide association meta-analysis of Parkinson’s disease. Nat Genet 2024; 56: 27–36.

6 Nalls MA, Blauwendraat C, Vallerga CL, et al. Identification of novel risk loci, causal insights, and heritable risk for Parkinson’s disease: a meta-analysis of genome-wide association studies. Lancet Neurol 2019; 18: 1091–102.

7 Loesch DP, Horimoto ARVR, Heilbron K, et al. Characterizing the Genetic Architecture of Parkinson’s Disease in Latinos. Ann Neurol 2021; 90: 353–65.

8 Leal TP, Waldo E, Duarte-Zambrano F, et al. Genotype-phenotype association study conducted on LARGE-PD reveals novel loci associated with Parkinson’s Disease. medRxiv. 2025; published online July 18. DOI:10.1101/2025.07.18.25331793.

9 Elsayed I, Martinez-Carrasco A, Cornejo-Olivas M, Bandres-Ciga S. Mapping the Diverse and Inclusive Future of Parkinson’s Disease Genetics and Its Widespread Impact. Genes 2021; 12. DOI:10.3390/genes12111681.

10 Schumacher-Schuh AF, Bieger A, Okunoye O, et al. Underrepresented Populations in Parkinson’s Genetics Research: Current Landscape and Future Directions. Mov Disord 2022; 37: 1593–604.

11 Okubadejo N, Britton A, Crews C, et al. Analysis of Nigerians with apparently sporadic Parkinson disease for mutations in LRRK2, PRKN and ATXN3. PLoS One 2008; 3: e3421.

12 Cilia R, Sironi F, Akpalu A, et al. Screening LRRK2 gene mutations in patients with Parkinson’s disease in Ghana. J Neurol 2012; 259: 569–70.

13 Okubadejo NU, Rizig M, Ojo OO, et al. Leucine rich repeat kinase 2 (LRRK2) GLY2019SER mutation is absent in a second cohort of Nigerian Africans with Parkinson disease. PLoS One 2018; 13: e0207984.

14 Yonova-Doing E, Atadzhanov M, Quadri M, et al. Analysis of LRRK2, SNCA, Parkin, PINK1, and DJ-1 in Zambian patients with Parkinson’s disease. Parkinsonism Relat Disord 2012; 18: 567–71.

15 Safiri S, Noori M, Nejadghaderi SA, et al. The burden of Parkinson’s disease in the Middle East and North Africa region, 1990-2019: results from the global burden of disease study 2019. BMC Public Health 2023; 23: 107.

16 Ojo OO, Abubakar SA, Iwuozo EU, et al. The Nigeria Parkinson Disease Registry: Process, Profile, and Prospects of a Collaborative Project. Mov Disord 2020; 35. DOI:10.1002/mds.28123.

17 Ojo OO, Wahab KW, Bello AH, et al. A Cross-Sectional Comprehensive Assessment of the Profile and Burden of Non-motor Symptoms in Relation to Motor Phenotype in the Nigeria Parkinson Disease Registry Cohort. Mov Disord Clin Pract 2021; 8: 1206–15.

18 Khedr EM, Fawi G, Abbas MAA, et al. Prevalence of Parkinsonism and Parkinson’s disease in Qena governorate/Egypt: a cross-sectional community-based survey. Neurol Res 2015; 37: 607–18.

19 El-Tallawy HN, Farghaly WM, Shehata GA, et al. Prevalence of Parkinson’s disease and other types of Parkinsonism in Al Kharga district, Egypt. Neuropsychiatr Dis Treat 2013; 9: 1821–6.

20 Zirra A, Rao SC, Bestwick J, et al. Gender Differences in the Prevalence of Parkinson’s Disease. Mov Disord Clin Pract 2023; 10: 86–93.

21 Singleton A, Blauwendraat C, Morris HR, et al. Parkinson’s disease: emerging opportunities through global collaboration. Lancet (London, England) 2025; published online Oct 9. DOI:10.1016/S0140-6736(25)01910-5.

22 Lim SY, Tan AH, Ahmad-Annuar A, et al. Uncovering the genetic basis of Parkinson’s disease globally: from discoveries to the clinic. The Lancet Neurology 2024; 23. DOI:10.1016/S1474-4422(24)00378-8.

23 GP2: The Global Parkinson’s Genetics Program. Movement disorders: official journal of the Movement Disorder Society 2021; 36. DOI:10.1002/mds.28494.

24 Junker J, Lange LM, Vollstedt EJ, et al. Team Science Approaches to Unravel Monogenic Parkinson’s Disease on a Global Scale. Movement disorders: official journal of the Movement Disorder Society 2024; 39. DOI:10.1002/mds.29925.

25 Chahine LM, Louie N, Solle J, et al. The Black and African American Connections to Parkinson’s Disease (BLAAC PD) study protocol. BMC neurology 2024; 24. DOI:10.1186/s12883-024-03914-7.

26 Lange LM, Fang ZH, Makarious MB, et al. The Global Landscape of Genetic Variation in Parkinson’s disease: Multi-Ancestry Insights into Established Disease Genes and their Translational Relevance. medRxiv: the preprint server for health sciences. 2025; published online July 11. DOI:10.1101/2025.07.08.25330815.

27 Ben-Joseph A, Marshall CR, Lees AJ, Noyce AJ. Ethnic Variation in the Manifestation of Parkinson’s Disease: A Narrative Review. Journal of Parkinson’s disease 2020; 10. DOI:10.3233/JPD-191763.

28 Schlebusch CM, Jakobsson M. Tales of Human Migration, Admixture, and Selection in Africa. Annu Rev Genomics Hum Genet 2018; 19: 405–28.

29 Choudhury A, Aron S, Botigué LR, et al. High-depth African genomes inform human migration and health. Nature 2020; 586: 741–8.

30 Rizig M, Bandres-Ciga S, Makarious MB, et al. Identification of genetic risk loci and causal insights associated with Parkinson’s disease in African and African admixed populations: a genome-wide association study. Lancet Neurol 2023; 22: 1015–25.

31 Álvarez Jerez P, Wild Crea P, Ramos DM, et al. African ancestry neurodegeneration risk variant disrupts an intronic branchpoint in GBA1. Nat Struct Mol Biol 2024; 31: 1955–63.

32 Ojo OO, Bandres-Ciga S, Makarious MB, et al. GBA1 rs3115534 Is Associated with REM Sleep Behavior Disorder in Parkinson’s Disease in Nigerians. Mov Disord 2024; 39: 728–33.

33 Hughes AJ, Daniel SE, Kilford L, Lees AJ. Accuracy of clinical diagnosis of idiopathic Parkinson’s disease: a clinico-pathological study of 100 cases. J Neurol Neurosurg Psychiatry 1992; 55: 181–4.

34 Verma A, Huffman JE, Rodriguez A, et al. Diversity and scale: Genetic architecture of 2068 traits in the VA Million Veteran Program. Science 2024; 385: eadj1182.

35 Bandres-Ciga S, Faghri F, Majounie E, et al. NeuroBooster Array: A Genome-Wide Genotyping Platform to Study Neurological Disorders Across Diverse Populations. medRxiv 2023; published online Nov 14. DOI:10.1101/2023.11.06.23298176.

36 Koretsky MJ, Alvarado C, Makarious MB, et al. Genetic risk factor clustering within and across neurodegenerative diseases. bioRxiv. 2022; published online Dec 3. DOI:10.1101/2022.12.01.22282945.

37 1000 Genomes Project Consortium, Auton A, Brooks LD, et al. A global reference for human genetic variation. Nature 2015; 526: 68–74.

38 Siva N. 1000 Genomes project. Nat Biotechnol 2008; 26: 256.

39 Bray SM, Mulle JG, Dodd AF, Pulver AE, Wooding S, Warren ST. Signatures of founder effects, admixture, and selection in the Ashkenazi Jewish population. Proc Natl Acad Sci U S A 2010; 107: 16222–7.

40 Taliun D, Harris DN, Kessler MD, et al. Sequencing of 53,831 diverse genomes from the NHLBI TOPMed Program. Nature 2021; 590: 290–9.

41 Das S, Forer L, Schönherr S, et al. Next-generation genotype imputation service and methods. Nat Genet 2016; 48: 1284–7.

42 Fuchsberger C, Abecasis GR, Hinds DA. minimac2: faster genotype imputation. Bioinformatics 2015; 31: 782–4.

43 Gagliano Taliun SA, VandeHaar P, Boughton AP, et al. Exploring and visualizing large-scale genetic associations by using PheWeb. Nat Genet 2020; 52: 550–2.

44 Chang CC, Chow CC, Tellier LC, Vattikuti S, Purcell SM, Lee JJ. Second-generation PLINK: rising to the challenge of larger and richer datasets. Gigascience 2015; 4. DOI:10.1186/s13742-015-0047-8.

45 Purcell S, Neale B, Todd-Brown K, et al. PLINK: a tool set for whole-genome association and population-based linkage analyses. Am J Hum Genet 2007; 81: 559–75.

46 He Y, Koido M, Shimmori Y, Kamatani Y. 2023; published online May 1. DOI:10.51094/jxiv.370.

47 Nalls MA, Pankratz N, Lill CM, et al. Large-scale meta-analysis of genome-wide association data identifies six new risk loci for Parkinson’s disease. Nat Genet 2014; 46: 989–93.

48 Reczek D, Schwake M, Schröder J, et al. LIMP-2 is a receptor for lysosomal mannose-6-phosphate-independent targeting of beta-glucocerebrosidase. Cell 2007; 131: 770–83.

49 Tan E-K, Peng R, Teo Y-Y, et al. Multiple LRRK2 variants modulate risk of Parkinson disease: a Chinese multicenter study. Hum Mutat 2010; 31: 561–8.

50 Taylor M, Alessi DR. Advances in elucidating the function of leucine-rich repeat protein kinase-2 in normal cells and Parkinson’s disease. Curr Opin Cell Biol 2020; 63: 102–13.

51 Vitte J, Traver S, Maués De Paula A, et al. Leucine-rich repeat kinase 2 is associated with the endoplasmic reticulum in dopaminergic neurons and accumulates in the core of Lewy bodies in Parkinson disease. J Neuropathol Exp Neurol 2010; 69: 959–72.

52 Dehay B, Bourdenx M, Gorry P, et al. Targeting α-synuclein for treatment of Parkinson’s disease: mechanistic and therapeutic considerations. Lancet Neurol 2015; 14: 855–66.

53 Yaribash S, Mohammadi K, Sani MA. Alpha-Synuclein Pathophysiology in Neurodegenerative Disorders: A Review Focusing on Molecular Mechanisms and Treatment Advances in Parkinson’s Disease. Cell Mol Neurobiol 2025; 45: 30.

54 Calabresi P, Mechelli A, Natale G, Volpicelli-Daley L, Di Lazzaro G, Ghiglieri V. Alpha-synuclein in Parkinson’s disease and other synucleinopathies: from overt neurodegeneration back to early synaptic dysfunction. Cell Death Dis 2023; 14: 176.

55 Sun W, Schulte C, Gasser T, Tan M, the Global Parkinson’s Genetic Program (GP2). TMEM175, SCARB2 and CTSB associations with Parkinson’s disease risk across populations. medRxiv. 2025; published online Aug 24. DOI:10.1101/2025.08.17.25333823.

56 Ross OA, Soto-Ortolaza AI, Heckman MG, et al. Association of LRRK2 exonic variants with susceptibility to Parkinson’s disease: a case-control study. Lancet Neurol 2011; 10: 898–908.

57 Bardien S, Lesage S, Brice A, Carr J. Genetic characteristics of leucine-rich repeat kinase 2 (LRRK2) associated Parkinson’s disease. Parkinsonism Relat Disord 2011; 17: 501–8.

58 Weindel CG, Bell SL, Vail KJ, West KO, Patrick KL, Watson RO. LRRK2 maintains mitochondrial homeostasis and regulates innate immune responses to Mycobacterium tuberculosis. 2020; published online Feb 14. DOI:10.7554/eLife.51071.

59 Tian Y, Lv J, Su Z, et al. LRRK2 plays essential roles in maintaining lung homeostasis and preventing the development of pulmonary fibrosis. Proceedings of the National Academy of Sciences 2021; 118: e2106685118.

